# Sequencing of 19,219 exomes identifies a low-frequency variant in *FKBP5* promoter predisposing to high myopia in a Han Chinese population

**DOI:** 10.1101/2022.09.06.22279641

**Authors:** Jianzhong Su, Jian Yuan, Liangde Xu, Shilai Xing, Mengru Sun, Yinghao Yao, Yunlong Ma, Fukun Chen, Longda Jiang, Kai Li, Xiangyi Yu, Zhengbo Xue, Yaru Zhang, Dandan Fan, Ji Zhang, Hui Liu, Xinting Liu, Guosi Zhang, Hong Wang, Meng Zhou, Fan Lyu, Gang An, Xiaoguang Yu, Myopia Associated Genetics and Intervention Consortium, Yuanchao Xue, Jian Yang, Jia Qu

**Affiliations:** School of Ophthalmology & Optometry and Eye Hospital, Wenzhou Medical University, Wenzhou, 325027, China; National Clinical Research Center for Ocular Disease, Wenzhou, 325027, China; Oujiang Laboratory, Zhejiang Lab for Regenerative Medicine, Vision and Brain Health, Wenzhou 325101, Zhejiang, China; Wenzhou Institute, University of Chinese Academy of Sciences, Wenzhou, 325011, China; Institute of PSI Genomics, Wenzhou, 325024, China; Key Laboratory of RNA Biology, Institute of Biophysics, Chinese Academy of Sciences, Beijing, 100190, China; School of Life Sciences, Westlake University, Hangzhou, Zhejiang 310030, China; Westlake Laboratory of Life Sciences and Biomedicine, Hangzhou, Zhejiang 310024, China

**Author notes:** To whom correspondence should be addressed. Jianzhong Su;, Yuanchao Xue;, Jian Yang;, Jia Qu. These authors contributed equally (Jianzhong Su, Jian Yuan, Liangde Xu, Shilai Xing, Mengru Sun).

**Keywords:** high myopia, whole-exome sequencing, rare variants, exome-wide association study, burden test

## Abstract

High myopia (HM) is one of the leading causes of visual impairment and blindness worldwide. Here, we report a whole-exome sequencing (WES) study in 9,613 HM cases and 9,606 controls of Han Chinese ancestry to pinpoint HM-associated risk variants. Single-variant association analysis identified three novel genetic loci associated with HM, including an East Asian ancestry-specific low-frequency variant (rs533280354) in *FKBP5*. Multi-ancestry meta-analysis with WES data of 2,696 HM cases and 7,186 controls of European ancestry from the UK Biobank discerned a novel European ancestry-specific rare variant in *FOLH1*. Functional experiments revealed a mechanism whereby a single G to A transition at rs533280354 disrupted the binding of transcription activator KLF15 to the promoter of *FKBP5*, resulting in decreased transcription of *FKBP5*. Furthermore, burden tests showed a significant excess of rare protein-truncating variants among HM cases involved in retinal blood vessel morphogenesis and neurotransmitter transport.

## Introduction

High myopia (HM) is generally defined as myopia with a spherical equivalent refractive error of less than or equal to -6.0 diopters (D). HM is associated with increased risk of serious ocular complications, most notably retinal degeneration or even detachment, which can cause severe vision impairment or blindness (Morgan et al., 2012; Saw et al., 2005). The prevalence of HM among schoolchildren was reported to be 2%-2.7% worldwide (Holden et al., 2016; Wong and Saw, 2016), with a great excess in East Asian populations (4.5%-21.6%) (Jung et al., 2012; Lin et al., 2004; Xu et al., 2021). Similar to many other common diseases, myopia has a complex etiology influenced by the interplay of genetic and environmental factors (Wojciechowski, 2011). Twin and family studies have demonstrated a high heritability for HM, which is estimated to be ∼90% (Guggenheim et al., 2000; Lopes et al., 2009).

Over the past decade, genome-wide association studies (GWAS) for refractive error or myopia have revealed more than 400 association loci in samples of predominantly European ancestry, but these loci account for a limited fraction of the heritability for refractive error (Hysi et al., 2020; Meguro et al., 2020; Tedja et al., 2018), likely because of small effects not discovered due to insufficient statistical power and causal variants, especially those of low allele frequency, not well captured in conventional GWAS. Compared to SNP arrays, whole-exome sequencing (WES) provides further opportunities to investigate rare coding variants not well tagged by array SNPs or imputation. Previous trio and family-based exome sequencing studies of individuals with monogenetic forms of HM have identified several rare coding mutations in known myopia genes, such as *SCO2* (Tran-Viet et al., 2013), *BSG* (Jin et al., 2017), *CCDC102B* (Hosoda et al., 2018), and *LRPAP1* (Aldahmesh et al., 2013). Such mutations, however, only explain a small fraction of HM cases in the population.

In this study, to conduct an unbiased screening of protein-coding variants associated with HM and better understand the genetic architecture of HM, we performed a WES study in 9,613 HM cases and 9,606 controls (aged from 6 to 18) of Han Chinese ancestry, the largest study of this kind. Exome-wide association analysis identified four loci significantly associated with HM after correcting for multiple tests. Among these loci, a previously unknown East Asian-specific risk variant rs533280354 in the *FKBP5* promoter was detected, which disrupted the binding site with the transcription factor KLF15, leading to altered *FKBP5* gene expression. In a burden test of rare variants, we uncovered a polygenic burden primarily arising from rare disruptive mutations distributed across many genes, particularly those involved in retinal blood vessel morphogenesis and photoreceptor-mediated neurotransmitter transduction. Taken together, these data suggest that population-based WES can discover functional risk alleles and provide important clues to elucidate the etiology of high myopia.

## RESULTS

### Dataset overview

A total of 9,852 HM and 11,375 controls from the Myopia Associated Genetics and Intervention Consortium (MAGIC) project were included in the WES study (STAR Methods). After removing samples showing poor sequencing quality or ambiguous sex status, population outliers identified by principal component analysis (PCA), one of each pair of closely related individuals, and control samples showing ancestral mismatch with cases, we retained 9,613 cases and 9,606 controls (STAR Methods; Table S1; Figure S1-S4). After quality control, a total of 3.37 million variants, including 1.11 million nonsynonymous variants and 169,383 insertions and deletions (indels), were retained for analysis (STAR Methods; Table S2; Figure S5-S7). Here we define variants with MAF > 5%, 0.5% < MAF < 5%, and MAF < 0.5% as common (1.5%), low-frequency (1.2%) and rare (97.3%) variants, respectively (Table S3). With the large sample size, our study has 80% power to detect modest effects occurring at a common (MAF=5%) with odds ratio [OR] of 1.23 or low frequency (MAF=0.5%) variant with OR of 1.77 (Figure S8). To discern HM-associated common, low-frequency, and rare variants, we performed multiple analyses (Figure 1A): a variant-level exome-wide association study (ExWAS), a meta-analysis of ExWAS between East Asian and European populations, and a gene-level rare variant association study (RVAS) analysis.

**Figure 1.**
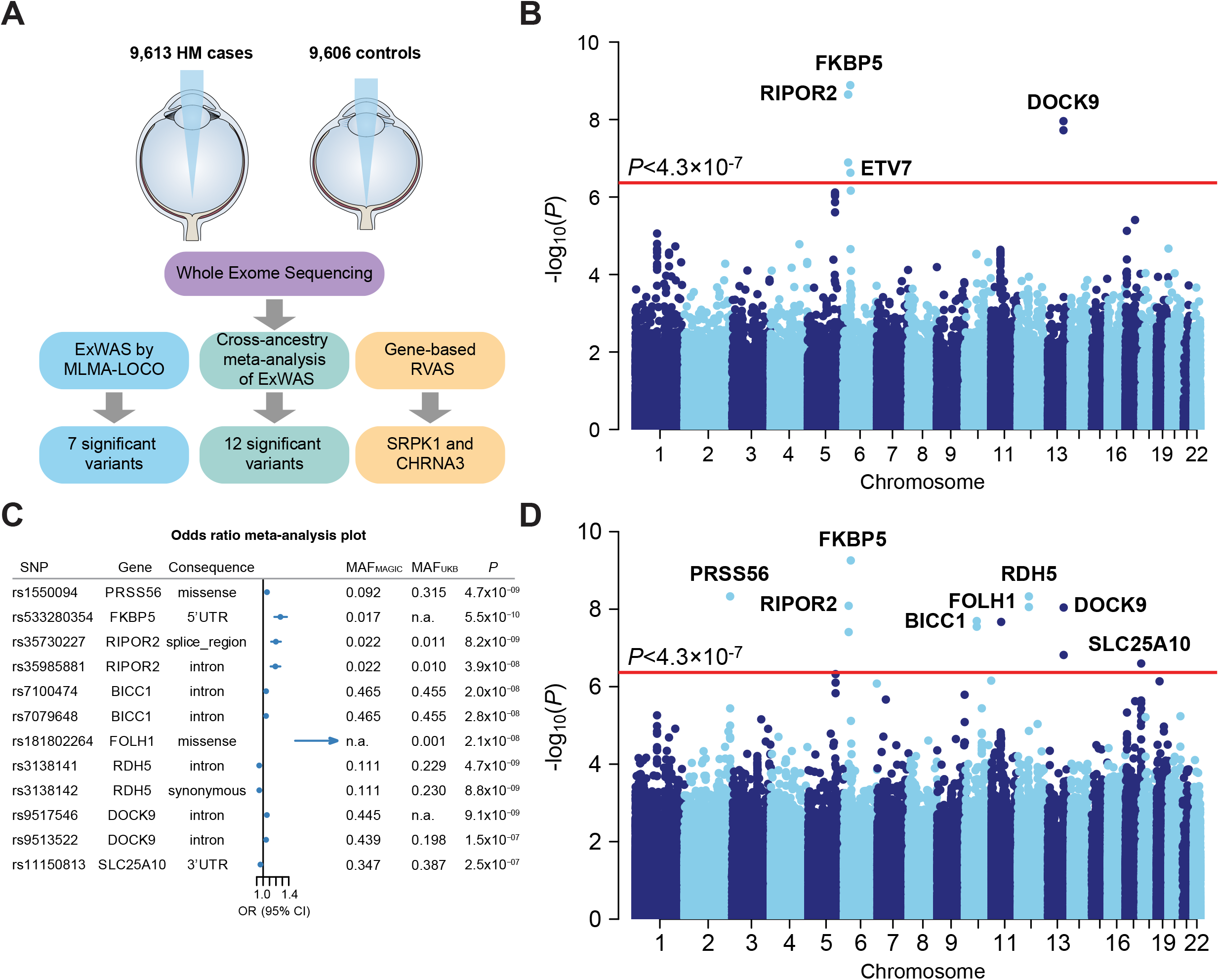
Study flowchart and variants ascertainment of exome-wide association analysis. (A) Overview of study design to identify and characterize high myopia-associated risk loci. HM: high myopia, ExWAS: exome-wide association studies, MLMA-LOCO: mixed linear model association (leaving-one-chromosome-out), PTV: protein-truncating variant, D-Mis: damaging missense variant. (B) Manhattan plot of single-variant associations analyses was calculated using the two-sided MLMA-LOCO test in MAGIC. The plot shows -log_10_-transformed *P* values for all SNPs. Red line for exome-wide significance, *P* = 4.3 × 10^−7^; Blue line for genome-wide significance, *P* = 5 × 10^−8^. (C) Forest plot of associations of 12 single nucleotide polymorphisms identified by ExWAS for association with HM risk. Odds ratios (95% CI) are presented for the meta-analysis of MAGIC and UK biobank data sets. (D) Manhattan plot of the ExWAS meta-analysis for HM in the combined analysis. The plot shows -log_10_-transformed *P* values for all SNPs. Red line, *P* = 4.3 × 10^−7^; Blue line, *P* = 5 × 10^−8^.

### Single variant-based exome-wide association analysis

We tested all 3.37 million variants each individually for association with HM (hereinafter referred to as ExWAS) using a linear mixed model-based association analysis method (MLMA-LOCO) that can account for population structure and relatedness (STAR Methods) (Yang et al., 2014). After adjusting the MLMA-LOCO test statistics by a genomic inflation factor of 1.57 (to correct for potential confounding factors not captured by the linear mixed model), we identified seven variants at four loci exceeding the exome-wide significance level of *P* < 4.3 × 10^−7^ (Sveinbjornsson et al., 2016) (Table 1; Figure 1B; Figure S9-S11). These include common variants at the known *DOCK9* gene locus (lead variant: rs9513522, MAF = 43%, *P* = 1.10 × 10^−8^), which is considered as a clinical hallmark of HM due to its association with irregular astigmatism and corneal ectasia (Gordon-Shaag et al., 2015), and low-frequency variants at three previously unknown loci (i.e., *FKBP5, RIPOR2* and *ETV7*). Specifically, we identified three low-frequency variants associated with HM in the intron region of *RIPOR2* (lead variants: rs35985881 and rs35730227, both with *P* = 2.26 × 10^−9^) and a low-frequency synonymous variant in *ETV7* (rs35985881, *P* = 2.36 × 10^−7^). Notably, we found a likely pathogenic variant (rs533280354, CADD = 16.47, *P* = 1.3 × 10^− 9^) located at the 5’UTR of *FKBP5*, which encodes a cis-trans prolyl isomerase that binds to the immunosuppressants FK506 and rapamycin and to a co-chaperone of Hsp90 which regulates glucocorticoid receptor (GR) sensitivity. The risk allele of this variant was much more frequent in East Asian populations (MAF=2.7%) than European (MAF=0.062%) or American populations (MAF=0.0%) according to the gnomAD data (Table S4). Consistently, it is a low-frequency in our cohort (MAF = 1.6%), with a relatively large effect size (allelic OR = 2.2 [1.77-2.73]) (Table 1).

**Table 1.**
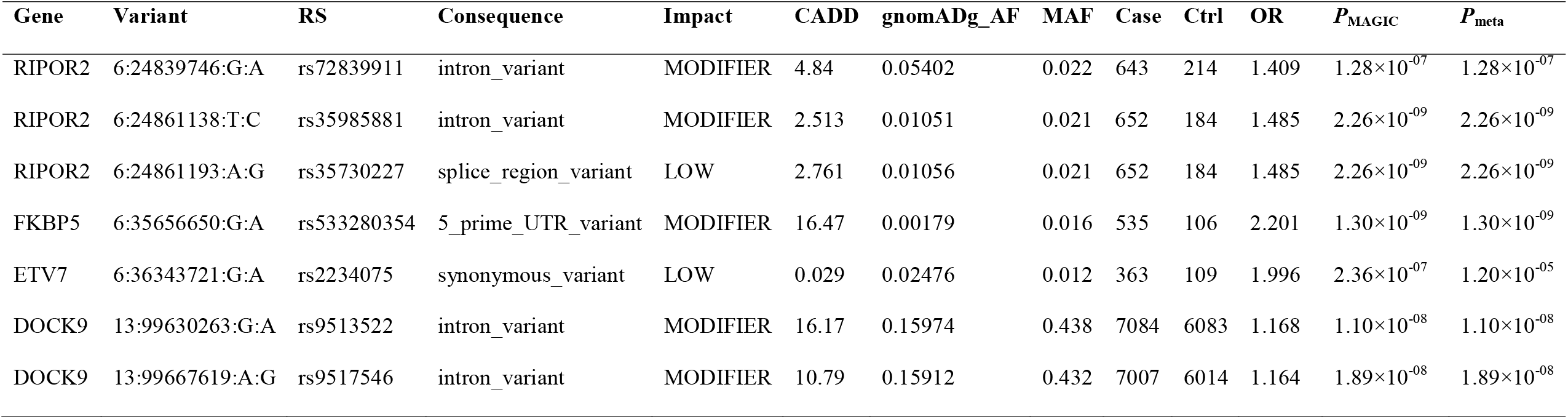
The most significant single-variant associations from exome-sequencing analysis in MAGIC.

| Gene | Variant | RS | Consequence | Impact | CADD | gnomADg_AF | MAF | Case | Ctrl | OR | $P_{\text{MAGIC}}$ | $P_{\text{meta}}$ |
| --- | --- | --- | --- | --- | --- | --- | --- | --- | --- | --- | --- | --- |
| RIPOR2 | 6:24839746:G:A | rs72839911 | intron_variant | MODIFIER | 4.84 | 0.05402 | 0.022 | 643 | 214 | 1.409 | $1.28 \times 10^{-07}$ | $1.28 \times 10^{-07}$ |
| RIPOR2 | 6:24861138:T:C | rs35985881 | intron_variant | MODIFIER | 2.513 | 0.01051 | 0.021 | 652 | 184 | 1.485 | $2.26 \times 10^{-09}$ | $2.26 \times 10^{-09}$ |
| RIPOR2 | 6:24861193:A:G | rs35730227 | splice_region_variant | LOW | 2.761 | 0.01056 | 0.021 | 652 | 184 | 1.485 | $2.26 \times 10^{-09}$ | $2.26 \times 10^{-09}$ |
| FKBP5 | 6:35656650:G:A | rs533280354 | 5_prime_UTR_variant | MODIFIER | 16.47 | 0.00179 | 0.016 | 535 | 106 | 2.201 | $1.30 \times 10^{-09}$ | $1.30 \times 10^{-09}$ |
| ETV7 | 6:36343721:G:A | rs2234075 | synonymous_variant | LOW | 0.029 | 0.02476 | 0.012 | 363 | 109 | 1.996 | $2.36 \times 10^{-07}$ | $1.20 \times 10^{-05}$ |
| DOCK9 | 13:99630263:G:A | rs9513522 | intron_variant | MODIFIER | 16.17 | 0.15974 | 0.438 | 7084 | 6083 | 1.168 | $1.10 \times 10^{-08}$ | $1.10 \times 10^{-08}$ |
| DOCK9 | 13:99667619:A:G | rs9517546 | intron_variant | MODIFIER | 10.79 | 0.15912 | 0.432 | 7007 | 6014 | 1.164 | $1.89 \times 10^{-08}$ | $1.89 \times 10^{-08}$ |

### Cross-population ExWAS meta-analysis

To assess the transferability of our findings in European populations and boost the power of detecting the associations shared between populations, we performed an ExWAS analysis of 2,696 HM cases and 7,186 controls of European ancestry in the UK Biobank (UKB) after quality control, which identified 5 quasi-independent, exome-wide significant variants at 3 loci (Figure S12-S13), including two previously reported common-variant loci (*PRSS56* and *TSPAN10*) (Tedja et al., 2018), and an unknown rare variant association in *FOLH1* (rs181802264, MAF = 0.106%, OR= 10.24, *P* = 2.00×10^−08^). Although the sample sizes were too small to obtain a precise estimate of the exome-wide genetic overlap between the two populations (the estimate of genetic correlation from Popcorn was 0.56 with a standard error of 0.41) (Brown et al., 2016), we observed significant between-population correlation of allelic effects (i.e., logOR) for variants clumped with different cut-off *P-*values (Figure S14). We then meta-analyzed the UKB and our MAGIC ExWAS summary statistics using METAL (Willer et al., 2010), and identified 12 quasi-independent, exome-wide significant SNPs (*P* < 4.3 × 10^−7^; Figure 1C, Table S5). These sentinel variants were annotated into 8 distinct genomic regions on 7 chromosomes (Figure 1D, Table S6). Among the 8 loci, 6 could be identified in the single ancestry ExWAS including three loci in the MAGIC and three loci in the UKB. The two loci that could be identified only by cross-population meta-analysis were *RDH5* (rs3138141, *P*_*meta*_=4.69×10^−09^) and *BICC1* (rs7100474, *P*_*meta*_=2.02×10^−08^), both of which have been reported to be associated with HM previously (Table S6). Together, we identified three previously unreported association signals, involving low-frequency or rare variants at the 5’UTR of *FKBP5* (rs533280354, *P*_meta_=5.53×10^−10^), a splice region of *RIPOR2* (rs35730227, *P*_*meta*_=8.19×10^−09^), and the *FOLH1* locus (rs181802264, *P*_*meta*_=2.13×10^−08^) (Table S5). Among these variants, the Chinese ancestry-specific variant rs533280354 in *FKBP5* was not examined in the UKB and the European-specific variant rs181802264 in *FOLH1* was not examined in the MAGIC (Table S5).

### Functional validation of the *FKBP5* variant *in silico* and *in vitro*

We next performed functional experiments to investigate the genomic and chromatin characteristics of the 5’UTR of *FKBP5*, as it contained the Chinese ancestry-specific variant rs533280354 that showed the strongest association with HM in our cohort (Figure 1B). Motif mining revealed that this variant was in the binding site (“CCCGCCC”) of transcription factor KLF15 (Figure 2A). Strong KLF15 ChIP-seq, H3K27ac and ATAC-seq enrichments were detected in the *FKBP5*’s 5’UTR region encompassing rs533280354 in retina or macula cell lines according to published data (Figure 2B). Gene expression analysis of *KLF15* and *FKBP5* indicated significant positive correlations with Pearson correlation coefficients of 0.41 (*P* = 3.5 × 10^−5^) in retina and 0.60 (*P* = 3.5 × 10^−11^) in retinal pigment epithelium (Figure 2C; Figure S15). These results prompted us to speculate that the *KLF15*-mediated transcriptional activation of *FKBP5* may be disrupted by alteration of transcription factor binding site where rs533280354 resides.

**Figure 2.**
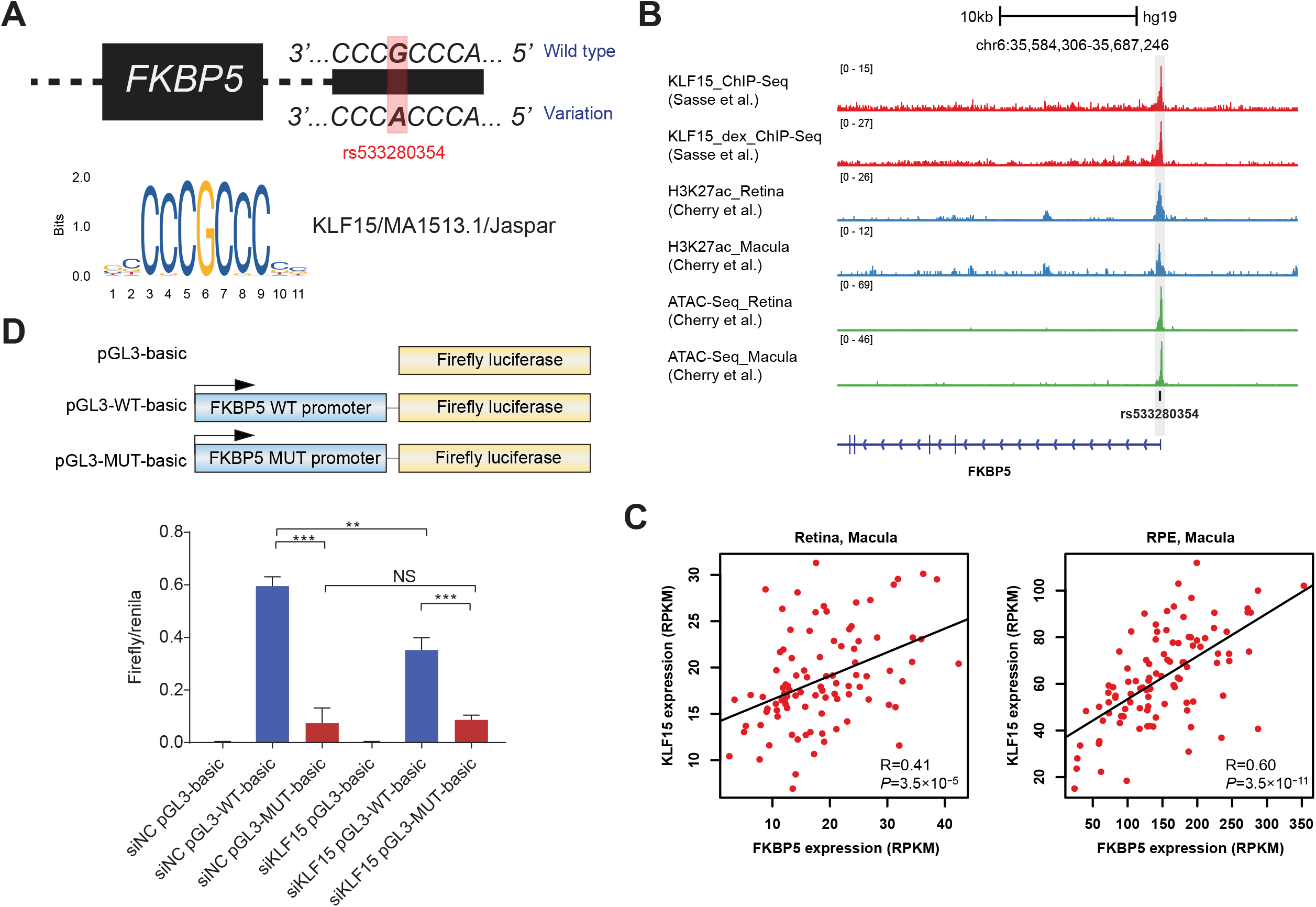
Functional dissection of exome-wide significant signal rs533280354. (A) *FKBP5* 5’UTR mutation alters transcription factor binding. The FKBP5 gene body is marked by a black box, with the intronic region represented by a dotted line. The wild-type and mutant DNA sequence for a small portion of the promoter region is indicated. The mutated *FKBP5* 5’UTR sequence is given, featuring a G > A mutation which destroy a consensus binding motif for a KLF15 transcription factor. (B) rs533280354 overlaps KLF15 ChIP-seq peak in HASM2 cells, H3K27ac peaks and an ATAC-seq peak in retina and macula, and is located in 5’UTR of FKBP5, transcribed right-to-left in the image. (C) mRNA expression correlation between *FKBP5* and KLF15 transcription factors in retina and RPE macula samples. (Insert) Pearson correlation coefficients were calculated. (D) Luciferase reporter assay of rs533280354. Outline of the luciferase reporter plasmid constructs used for transfection. The plasmids consisted of *FKBP5* promoter containing rs533280354 that were sub-cloned into the pGL3 vector. The ability of these plasmid constructs to enhance transcription in transfected HEK293 cells was measured by determination of cellular luciferase (luc) activity 24 hours after transfection. The luciferase activities of cells transfected with the susceptibility allele (A-allele) of rs12946510 were reduced compared to those transfected with the major allele (G-allele). siRNA-mediated knockdown of KLF15 also deregulates *FKBP5* expression in HEK293 cells. Three independent experiments were performed in each assay. The data in the figures represent averages ± standard deviation of triplicate assays in one experiment. \*\**P* < 0.01, \*\*\**P* < 0.001 (Student’s t test).

To examine whether the identified KLF15 binding motif is necessary for *FKBP5* activation, we cloned the 5’UTR of wild-type (WT, G allele in rs533280354) promoter sequence and mutant sequence (MUT, A allele in rs533280354) into the pGL3-basic luciferase vector (Figure 2D). We found that compared with pGL3-basic, the inserted WT promoter sequence exhibited strong luciferase activity in HEK293 cells, but the MUT promoter (with a single G-to-A transition) showed dramatically reduced transcriptional activation of the firefly luciferase gene (*P* = 5.5×10^−4^, two-tailed *t*-test; Figure 2D), indicating that rs533280354 is a causal variant directly influencing KLF15 binding to the promoter region of *FKBP5*.

To test this hypothesis, we used small interfering RNA (siRNA) to knock down KLF15 and then examined the luciferase activities of WT and MUT *FKBP5* promoters. We found that the knocking down KLF15 with siRNAs reduced the WT promoter by 40.9%, whereas MUT promoter was not changed (*P* = 0.0026, two-tailed t test; Figure 2D; Figure S16). Together, these data suggested a molecular mechanism whereby a single G-to-A transition at rs533280354 leads to decreased gene transcription by disrupting the KLF15 binding motif at the promoter region of *FKBP5*.

### Rare variants burden analysis for HM

Because single-variant analyses have limited power for detecting rare-variant associations, we next performed set-based analyses to aggregate the effects of multiple rare variants, here defined as variants with MAF less than 0.5% in our dataset, the 1000 Genomes, NHLBI Exome Sequencing Project (ESP), and gnomAD. We first evaluated the association of the burden of all rare protein-truncating variants (PTVs), damaging missense (D-Mis), benign missense, or synonymous variants with HM by three logistic models, and then dissect the burden test into those focusing on specific gene sets (STAR Methods, Table S7, Figure S17). In the burden test, each model used Firth-based logistic regression and incorporated some or all of the following covariates: sex, the first ten principal components (PCs) derived from common variants, and variant count (i.e., summation of rare synonymous, benign missense, damaging missense and PTVs) (STAR Methods, Figure S17). According to the relatively most conservative model which included all the covariates, we observed a significant enrichment of rare PTVs (OR = 1.04, *P* = 1.76 × 10^−10^) and D-Mis (OR = 1.03, *P* = 6.71 × 10^−18^) in HM cases relative to controls (Figure 3A). There was no evidence of excess burden in rare synonymous variants, which can be regarded as a negative control, suggesting that enrichment of deleterious pathogenic variants was unlikely to result from un-modeled population stratification or technical artifact. Next, we limited the burden test to rare variants in the known myopia-related genes obtained from the IMI - Myopia Genetics Report (Table S8) (MS et al., 2019). We found a pronounced difference in the number of PTVs, but not D-Mis or benign and synonymous variants, in HM individuals relative to controls (OR = 1.89, *P* = 3.0 × 10^−04^), suggesting a potentially unique role of PTVs in the known myopia genes in HM risk (Figure 3B). Besides, a significant enrichment of rare PTVs in genes related to retinal blood vessel morphogenesis (OR = 2.47, *P* = 3.19 × 10^−05^; Figure 3C) or neurotransmitter transport (OR = 2.07, *P* = 0.00016; Figure 3D) was detected, in line with the significant enrichment of common variant association signals in targets of angiogenesis and rod-and-cone bipolar synaptic neurotransmission shown in GWASs for refractive error (Hysi et al., 2020; Tedja et al., 2018).

**Figure 3.**
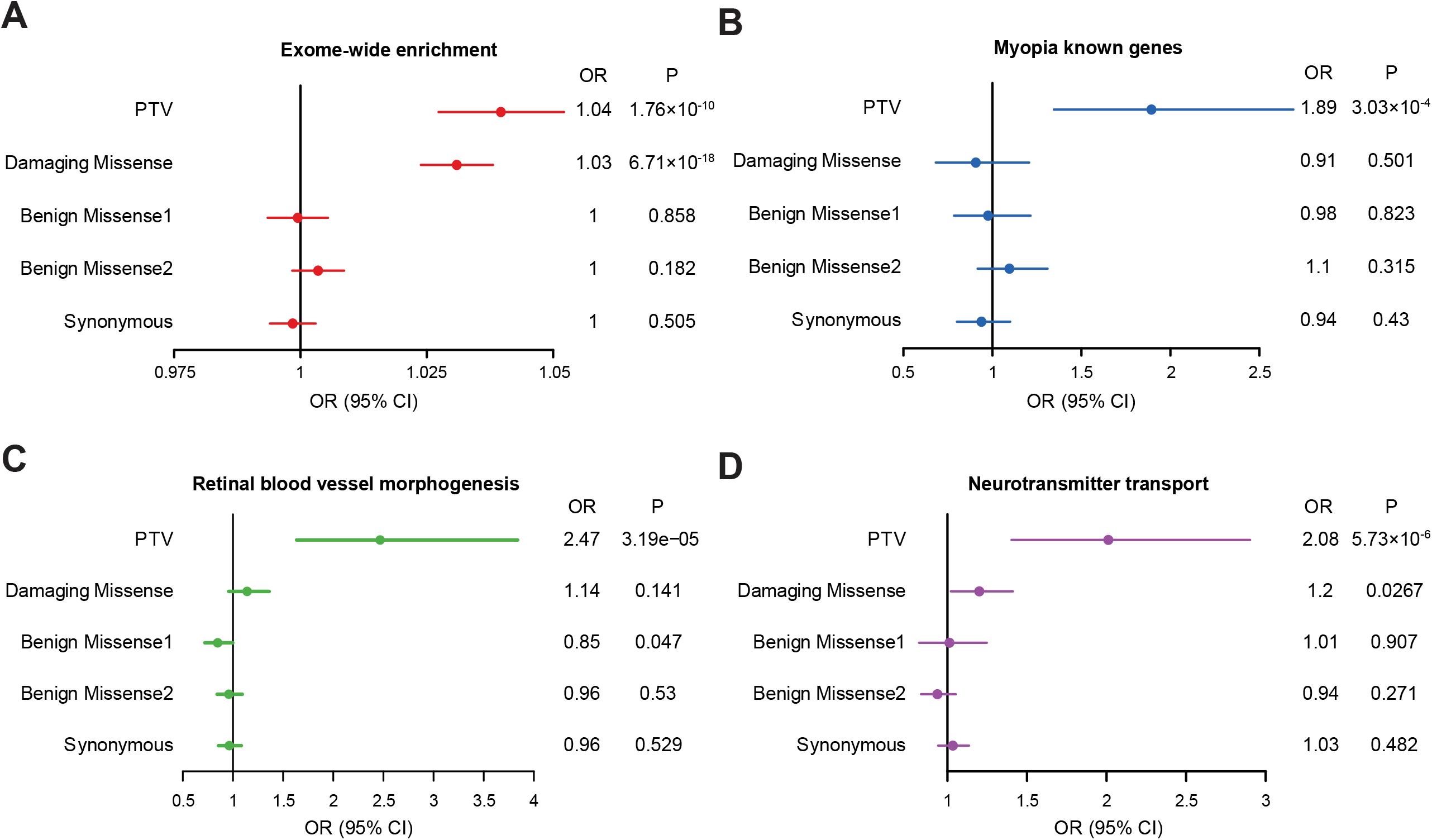
Exome-wide and gene-set burden analysis of PTVs and D-Mis in HM cases. (A) Exome-wide burden analysis of synonymous variants, benign missense variants, damaging missense variants and PTVs within rare variants (MAF<0.5%). Significance of association is displayed with the *P* values from Firth logistic regression test; errors bars indicates 95% confidence intervals (CIs) of the corresponding odds ratios. Multiple test correction *P* = 0.01. The graphs display the means and standard deviations. (B) Burden analysis of myopia-related GWAS genes only. Synonymous variants, benign missense variants, damaging missense variants and PTVs within rare variants are displayed. (C) Burden analysis of genes involved in retinal blood vessel morphogenesis. (D) Burden analysis of genes involved in neurotransmitter transport.

Biological and empirical gene sets can refine our understanding of the mechanisms underpinning association between rare variants and HM risk and help to derive potential biological hypotheses for subsequent investigations. To elucidate pathways enriched for rare variants in HM cases, collapsing analysis via Fisher’s exact test (Povysil et al., 2019) was performed for PTVs, D-Mis, benign missenses and synonymous variants to evaluate whether there is a significant difference between the counts of cases and controls who carry at least one qualifying variant. All gene sets derived from Gene Ontology (GO) categories, KEGG and REACTOME, as well as transcription factor targets from GSEA (10,587 gene sets). Results indicated that rare PTVs were significantly (*P*<0.05/10,587/5=9.44×10^−7^) enriched in biological processes related to Retinal blood vessel morphogenesis (GO:0061304; OR = 4.80, *P* = 2.04 × 10^−19^) and Neurotransmitter transport (GO:0006836; OR = 1.33, *P* = 2.47 × 10^−08^), as well as Regulation of TORC2 signaling (GO:1903939; OR = 28.06, *P* = 1.10 × 10^−07^), Vitamin biosynthetic process (GO:0009110; OR = 6.31, *P* = 1.17 × 10^−07^) and Extracellular matrix cell signaling (GO:0035426; OR = 5.46, *P* = 1.38 × 10^−19^), while rare D-mis were enriched in pathway of cellular response to light intensity (GO:0071484; OR = 3.92, *P* = 2.63 × 10^−09^), after correcting for multiple tests (Figure S18, Table S9-S10). The pathway collapsing analyses remained significant when tested against an empirical distribution generated by repeated sampling of the same number of length-matched genes at random 1000 times (Table S9-S10). No significant pathway enrichment was observed for benign missenses and synonymous variants, which can be considered as a negative control (Table S11-S12).

### Gene-based rare variant association analysis

We performed gene-based rare variant association analysis under the assumption that the detection power can be improved by aggregating the effects of multiple HM rare causal variants in a gene. The gene-based analysis was performed with five methods (i.e., Fisher’s exact test [FET], Burden, SKAT, SKAT-O and SAIGE-GENE) for robustness check. The test statistics from different methods showed high positive correlations (Figure S19), but only the SAIGE-GENE method (Zhou et al., 2020) had no evidence of inflation (Figure S20). After correcting for multiple tests, no gene reached the exome-wide significance level (*P* = 0.05/19574/5 = 5.11×10^−7^). However, the top two genes, *SRPK1* and *CHRNA3*, prioritized by SAIGE-GENE, showed strong deviations from the null hypothesis in the FET test (Figure 4; Figure S20) despite the overall inflation of the FET test statistics.

**Figure 4.**
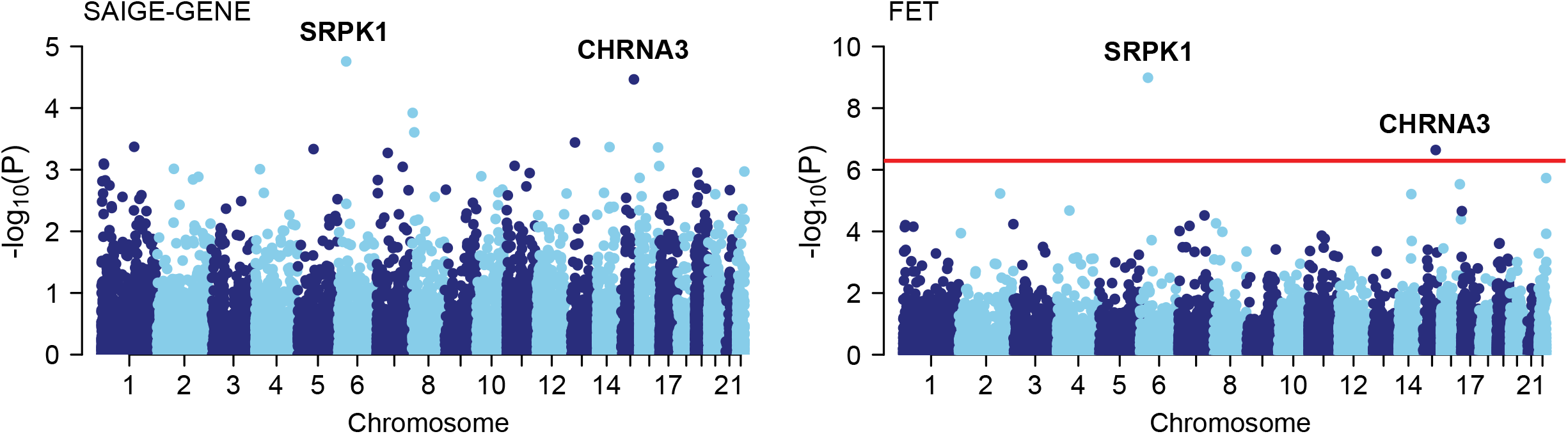
Gene discovery in the MAGIC Cohort. Manhattan plots of the gene-based burden analysis. An excess of rare deleterious (PTV and D-Mis) variants in HM cases within genes were tested using SAIGE-GENE and Fisher’s exact test [FET], Red line, *P* = 0.05/19574/5 = 5.11×10^−7^.

The top hit, *SRPK1* (the gene encoding serine/arginine protein kinase specific for the serine/arginine-rich domain family of splicing factors), has been reported to be involved in development of retinal blood vessels (Coultas et al., 2005; Gammons et al., 2013). It contained three PTVs (two frameshift variant: p.Ile192LeufsTer32 and p.Thr411ArgfsTer9, and a stop gained variant: p.Tyr365_Gln367delinsTer) and 7 D-Mis (p.Ala114Ser, p.Arg126Trp, p.Arg132Cys, p.Lys274Gln, p.Gly358Glu, p.Lys506Glu and p.Gly574Glu) in 122 of 9,613 HM cases and 12 of 9,606 controls (OR=10.28, GC corrected: *P*_SAIGE-GENE_ = 1.75 × 10^−05^, *P*_FET_ = 1.03 × 10^−09^; Figure 5A; Table S13). The gene *CHRNA3*, which encodes a subunit of the nicotinic acetylcholine receptor (AChR) that mediates fast synaptic transmission (Wang et al., 2002), harbored five rare PTVs (four frameshift variant: p.Leu17ArgfsTer37, p.Leu18AlafsTer17, p.Leu303AspfsTer115 and p.Glu477ValfsTer21, and a splice donor variant: c.377+2_377+5del), and 7 D-Mis (p.Arg110Cys, p.Ile249Leu, p.Leu253Met, p.Tyr264Phe, p.Gly270Ser, p.Thr284Met and p.Leu499Met) in 178 cases and 51 controls (OR=3.53, GC corrected: *P*_SAIGE-GENE_ = 3.43 × 10^−05^, *P*_FET_ = 2.28 × 10^−07^; Figure 5A; Table S13). The role of cholinergic receptors during myopia development was strengthened by the identification of previous GWAS of susceptibility loci for myopia on chromosome 15q25 including *CHRNA3* (Hysi et al., 2010).

**Figure 5.**
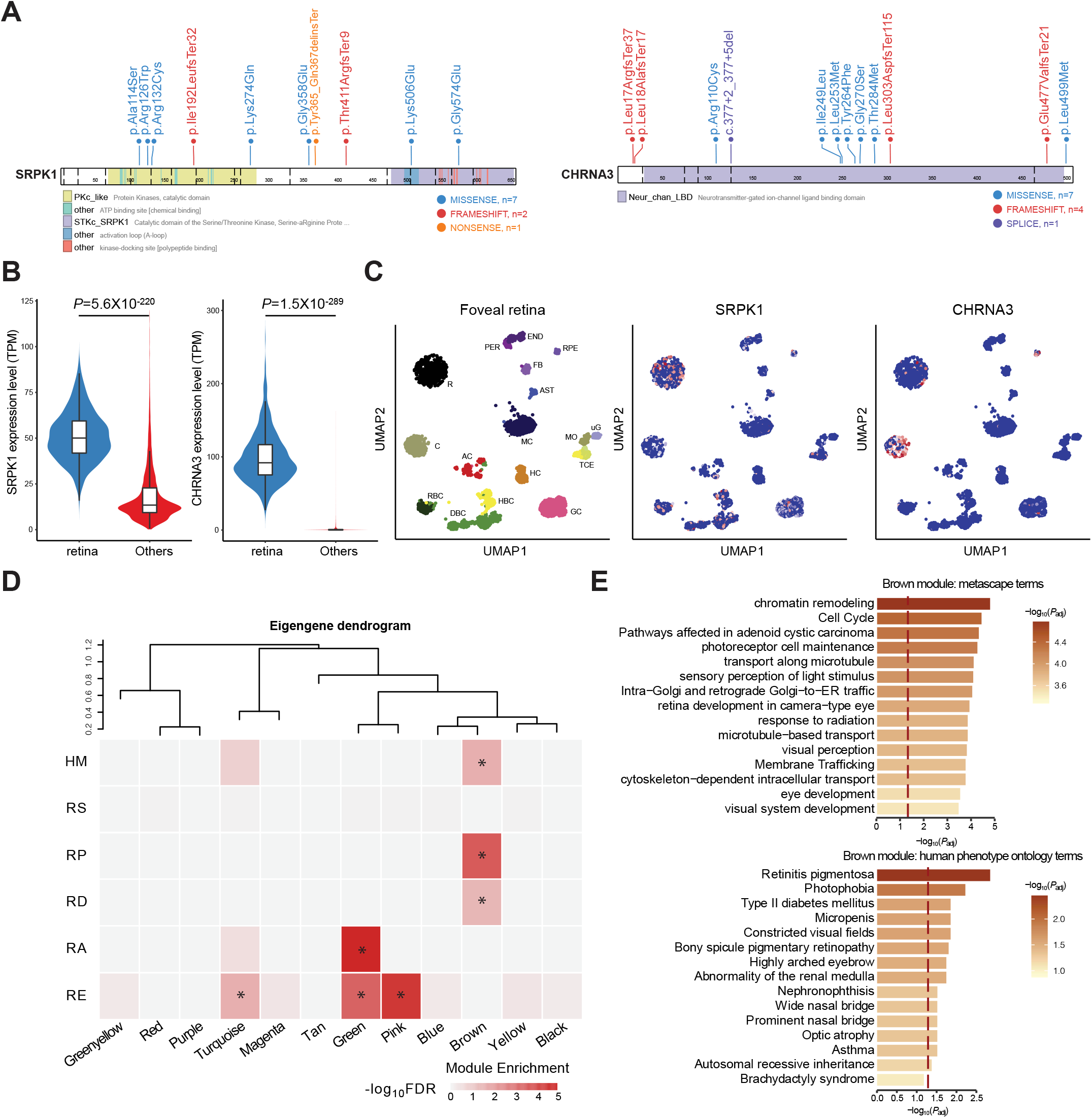
Functional categories and genetic characterization of HM-associated genes. (A) Mutation mapping of the two HM risk genes, *SRPK1* and *CHRNA3*, in relation to functional domains in each molecule. (B) Violin plots indicate the expression of risk genes associated with HM (*SRPK1* and *CHRNA3*) in retina and other tissues in GTEx. (C) Uniform manifold approximation and projection (UMAP) of all tissues single-cell data with cell colored based on the expression of *SRPK1* and *CHRNA3* genes for particular cell types. Gene expression levels are indicated by shades of red. (D) Enrichment analysis across weighted gene coexpression network analysis (WGCNA) modules of the human retina for genes with rare risk variation in HM, RP, RD, RA and RE. Only 12 modules are shown (labeled by color). Tiles are labeled with -log10(*P* value) and an asterisk represents statistically significant enrichment at the Bonferroni multiple-testing cutoff (α = 0.05/12 = 4.16 × 10^−3^). (E) Top 20 GO metascape terms and top 20 HP ontology terms enriched for the brown module. The x axis depicts -log (adjusted *P* value) and the dotted line represents the α = 0.05 significance threshold.

Based on bulk RNA sequencing (RNA-seq) data of 55 different tissues from the Genotype-Tissue Expression (GTEx) consortium (Battle et al., 2017), we found that *CHRNA3* and *SRPK1* were significantly over-expressed in retina than the other 54 tissues (Figure 5B). Cell-type specificity analysis of data from human peripheral and foveal cells (Cowan et al., 2020) showed that as a neurotransmitter synthesis and transport gene, *CHRNA3* was mainly expressed in numerous cone cells and RPE (Figure 5C; Figure S21-S22), suggesting that deleterious mutation in *CHRNA3* might affect rod-and-cone synaptic neurotransmission to RPE.

### HM risk genes converge in human retina coexpression network pertinent to other retinal disorders

Evidence from animal and preclinical studies have shown that retinal cell physiology could be disrupted by many myopia and refractive error risk variants (Gifford et al., 2019; Troilo et al., 2019). We selected the top 50 HM risk genes with pathogenic variants by SAIGE-GENE *P* values to test whether they converged in gene coexpression networks of retina (Ratnapriya et al., 2019; Yuan et al., 2021) and correlated with risk genes of other eye diseases (Figure S23). Notably, these HM risk genes converged in a single transcriptional network (‘brown’ module; FDR = 0.018; Figure 5D) which was also associated with retinal degeneration (RD) and retinitis pigmentosa (RP), but not with refractive error (RE) or refractive astigmatism (RA). The enrichment remained significant when tested against an empirical distribution generated by repeated sampling of 50 length-matched genes at random (*P* < 1.0×10^−3^; Figure S24).

Consistent with the functional enrichment analysis of HM risk genes, the top enriched GO biological process terms for the brown module included photoreceptor cell maintenance, sensory perception of light stimulus and retina development in camera-type eyes (Figure 5E, Table S14). The top enriched human phenotype (HP) ontology terms corresponded to ocular abnormalities, including ‘retinitis pigmentosa’ and ‘photophobia’ (Figure 5E, Table S15). These data suggest that mutations in candidate HM risk genes affect pathways that tend to be shared more with RD, RP, and other severe retinal disorders than ordinary refractive error or astigmatism.

## Discussion

HM is a leading cause of vision impairment and blindness. The global prevalence of HM is estimated to reach approximately 10% by 2050, affecting nearly 1 billion people (Holden et al., 2016). However, it remains a big challenge for the precise and efficient prevention and control of HM due to its heterogeneous etiology because of involvement of many genetic and environmental factors. To deepen the understanding of the genetic landscape as well as biological mechanisms of HM and give new insights into its potential relationship with other eye diseases, we assembled one of the largest cohort of schoolchildren with sporadic HM for WES. We identified significant low-frequency variant associations at three previously unknown loci in the exome-wide association analysis and association of the burden of rare PTVs or D-Mis with HM in the burden test, indicating an important role of rare and low-frequency variants in the predisposition to HM.

Both single- and cross-population ExWAS were carried out to identify ancestry-specific or - common HM risk variants. For example, we found a previously unknown Chinese-ancestry specific risk variant (rs533280354) in a binding site of the transcription factor KLF15 in the 5’UTR of *FKBP5*. The risk variant (G>A) conferred significantly lower transcriptional activity than the wild-type promoter sequence. In the Mouse Genome Informatics Database (MGI), the phenotypic effects of the mutations in *FKBP5* were the presence of corneal opacity, increased corneal thickness, and abnormal retina inner nuclear morphology (Eppig, 2017). *FKBP5* gene encodes a molecular cochaperone of the glucocorticoid receptor complex, which modulates intracellular glucocorticoid signaling (Binder, 2009; Zannas et al., 2016). When it is bound to the receptor complex, cortisol binds with lower affinity, and nuclear translocation of the glucocorticoid receptor (GR) becomes less efficient. Excessive glucocorticoids (GCs) or glucocorticoid receptor (GR) hypersensitivity may aggravate several underlying diseases, including myopia, which has been verified in an experimental lens-induced myopia animal model upon intraperitoneal injection of GCs (Ding et al., 2018; Zhang et al., 2021). Additionally, GCs have been shown to regulate inflammation immune response and inhibit growth of retinal blood and lymphatic vessels (Sulaiman et al., 2018). Therefore, the HM risk variant (A-allele) in *FKBP5* may induce glucocorticoid receptor sensitivity to GCs, which might increase the rate of axial elongation, affecting retinal blood vessel morphogenesis and promoting a myopic shift in refractive error. Besides, we also identified a previously unknow European-ancestry specific variant rs181802264 (p.Leu4Phe) in *FOLH1* with a large effect size (OR=11.39). *FOLH1* encodes a type II transmembrane glycoprotein and acts as a glutamate carboxypeptidase involved in excitatory neurotransmission process (Zink et al., 2020). This finding provides genetic support for the dysregulation of neurotransmission process as a possible mechanism of HM.

As mentioned above, we found from a burden test an overall enrichment of rare PTVs or D-mis. Although no single gene surpassed exome-wide significance in gene-based test, specific gene sets previously described to be relevant to myopia or retinal-related biological processes showed a discernable enrichment of rare variants associated with HM. Specifically, we observed a significant excess of rare PTVs in previously reported myopia-associated genes, including TORC2 signaling genes, extracellular matrix cell signaling genes, vitamin biosynthetic genes, and a large group of genes delineating retina blood vessel morphogenesis and neurotransmitter transports. Recently, a GWAS of HM in Asians reaffirmed the relevance of the nervous system to the pathogenesis of HM (Meguro et al., 2020), consistent with the evidence from several experimental models showing that the light-induced retina-to-sclera signaling pathway and rod-and-cone bipolar synaptic neurotransmission play key roles in refractive error or myopia development (Hysi et al., 2014; Tedja et al., 2018; Troilo et al., 2019; Verhoeven et al., 2013). However, the human pathobiological molecular drivers of HM genes in the retina-to-sclera signaling cascade or other potential pathways remain to be elucidated. Although there was no gene reached the exome-wide significance with the current sample size, our gene-based tests revealed several genes with suggestive evidence of association with HM, including *SRPK1* and *CHRNA3*, recapitulating known biological process involved in the neurotransmission and retina blood vessel morphogenesis. This reflects the highly polygenic genetic architecture of HM and suggests that discovering the majority of genes involved in HM risk will require large sample sizes.

To explore genetic relationships between HM and other retinopathy disorders (e.g., RD and RP), the overlap of risk genes between HM, RA, RE RD and RP were evaluated. Data demonstrated that HM, RD, and RP risk genes were enriched in the same co-expression network module, along with overlap of risk pathways between them. Convergence of HM, RD and RP risk genes in photoreceptor cell maintenance were discovered, perhaps accounting for the increased frequency of structural eye abnormalities in HM cases relative to individuals with low myopia. Additionally, consistent with a recent hypothesis (Hysi et al., 2020; Hysi et al., 2016), our results demonstrated that HM shows a genetic etiology distinct from that of common refractive error such as RE and RA.

Our study had some limitations. First, although 9,613 HM cases and 9,606 controls have been included in the WES study, the number of HM-associated risk genes discovered was limited, likely because the polygenic nature of the disease so that much larger sample sizes are required to unveil the remaining risk genes. Second, mechanistic insights into the newly identified HM risk genes and related pathways need further exploration by *in vivo* experiments in model organisms. The successful pursuit of these next steps will refine current heuristics for clinical decision making and give rise to therapeutic intervention for individuals with HM.

## Data Availability

Individual-level data are not publicly available due to ethical and legal restrictions related to the Wenzhou Medical University. Supporting data are available from the corresponding author upon reasonable request but access to data must be granted by the MAGIC committees. VCF files will be uploaded to Genome Variation Map (GVM) in BIG Data Center (http://bigd.big.ac.cn/gsa), Beijing Institute of Genomics (BIG), Chinese Academy of Sciences, with an accession number GVM000296. The datasets used and analyzed during the current study are available from the corresponding author on reasonable request.

## ACKNOWLEDGMENTS

We thank Dr. Zhenhui Chen, Dr. Yunlong Ma and Jiangfan Chen for constructive comments regarding this manuscript. We thank the Berry Genomics Co., Ltd for sequencing services. We thank Dr. Chaolong Wang for technical assistance. This work was supported by the National Natural Science Foundation of China (61871294, 82172882), Zhejiang Provincial Natural Science Foundation of China (LR19C060001), and the Scientific Research Foundation for Talents of Wenzhou Medical University (QTJ18023) to J. Su.

## CONSORTIA

The members of the Myopia Associated Genetics and Intervention Consortium (MAGIC) are Jianzhong Su, Jian Yuan, Liangde Xu, Shilai Xing, Mengru Sun, Yinghao Yao, Fukun Chen, Kai Li, Xiangyi Yu, Zhengbo Xue, Yaru Zhang, Ji Zhang, Hui Liu, Dandan Fan, Guosi Zhang, Hong Wang, Meng Zhou, Hao Chen, Fan Lyu, Gang An, Yuanchao Xue, Zhenhui Chen, Jian Yang, Jia Qu, Zhenhui Chen, Yunlong Ma, Yichun Xiong, Xinting Liu, Nan Wu, Jie Sun, Jinhua Bao, Liang Xu, Ling Li, Liang Ye, Jun Jiang, Xinjie Mao, Xinping Yu, Xiaoming Huang, Jingjing Xu, Miaomiao Li, Xuemei Zhang, Liang Hu, Zhuopao Zuo, Wanqing Jin, Jiawei Zhou, Yuwen Wang, Xue Li, Fang Hou, Yukuan Huang, Fei Qiu, Yijun Zhou, Na Gao, Xinyu Wang, Xinrui Shi, Yuchun Deng, Xiaoguang Yu, Yu Bai, Chenghao Li, Lu Chen, Ke Li, Lijun Dai, Xiangyi Yu, Peng Lin, Jingting Zhao, Congcong Yan, Siqi Bao, Zicheng Zhang, Fangjie Guo, Hongchen Han, Shen Wang, Haojun Sun, Siyi Jiang, Wei Dai, Hengte Kong, Xiaoyan Lu, Jing Li, Liansheng Li, Siyu Wang.

## AUTHOR CONTRIBUTIONS

The study was conceived, designed and supervised by J.Q., J.S., J.Y. and Y.X.. Analysis of data was performed by J.Y., J.S., J.Y., Y.M., S.X., L.X., Y.Y., F.X., L.J., Z.X, X.Y., J.Z. Y.Z. D.F. and H.W. The experiments were conducted by M.S. J.Y. and Y.X. Patient sample recruitment was conducted by J.Q., L.X., J.S., F. L., H.C., G.Z., H.L., M.Z., X.Y. and number of Myopia Associated Genetics and Intervention Consortium. DNA extraction and sequencing were carried out by S.X and G.X., The manuscript was written by J.Y., J.S. and S.X. with contributions from all other authors.

## DECLARATION OF INTERESTS

The authors declare no competing financial interests.

## STAR Methods

### KEY RESOURCES TABLE

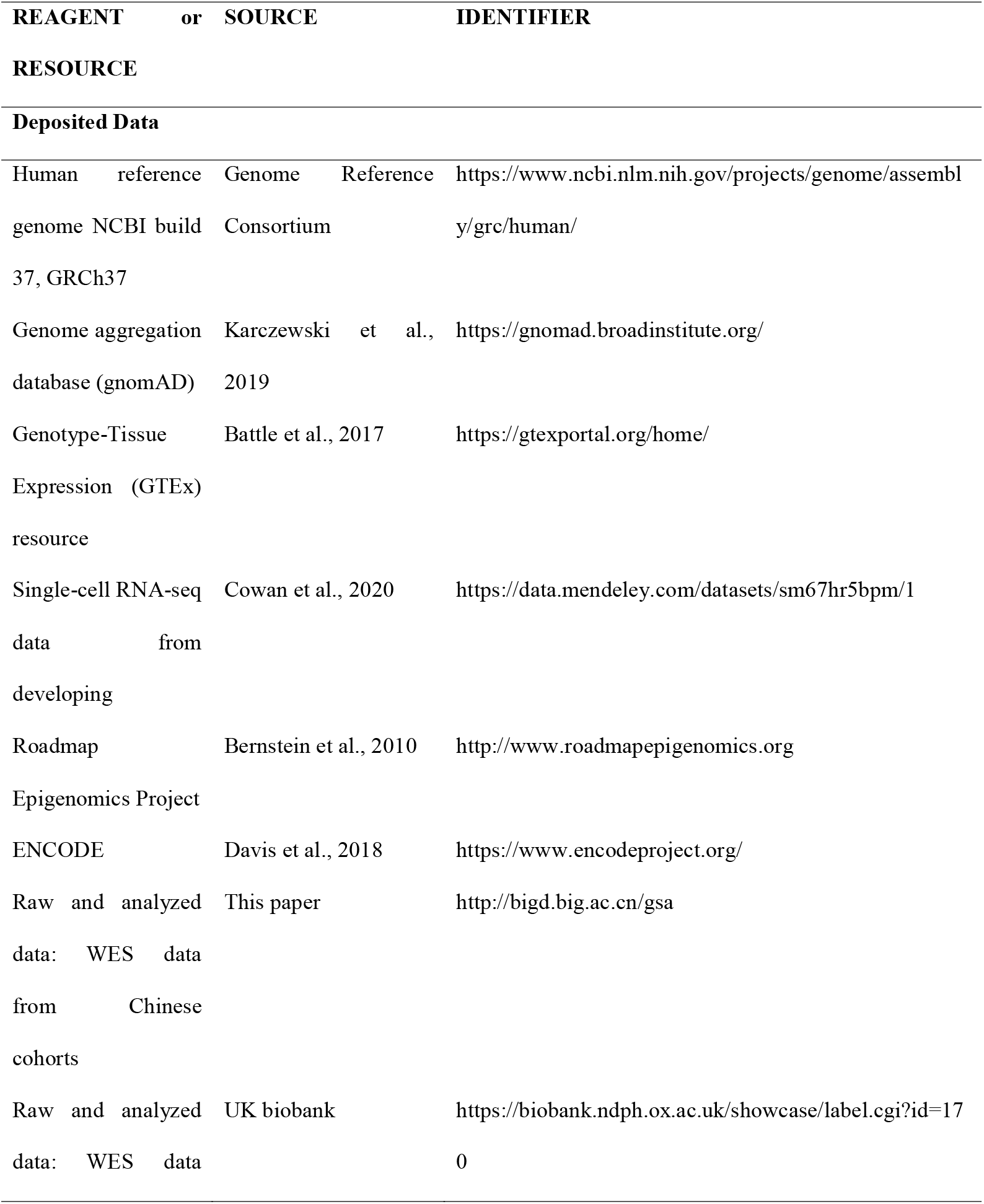
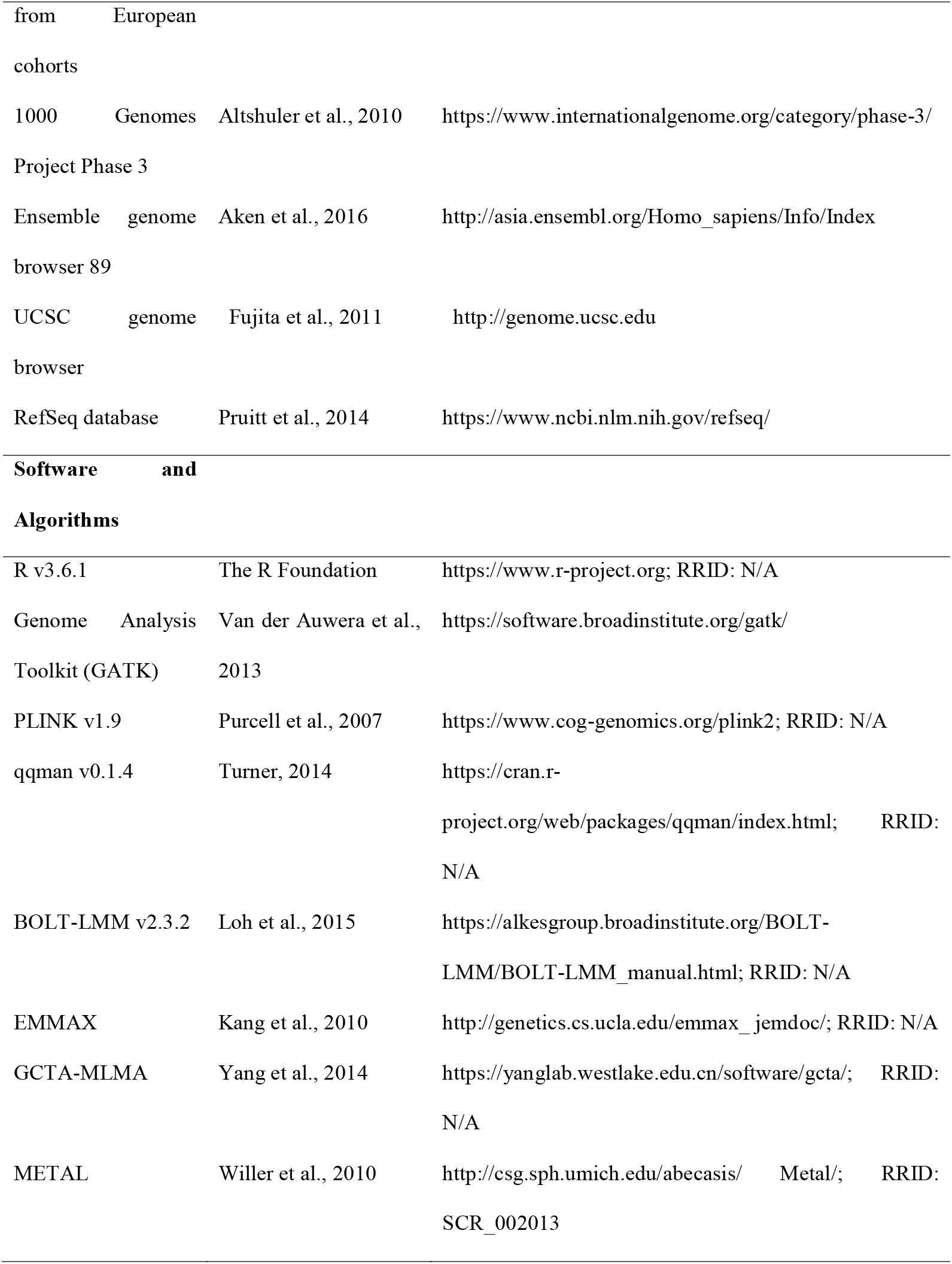
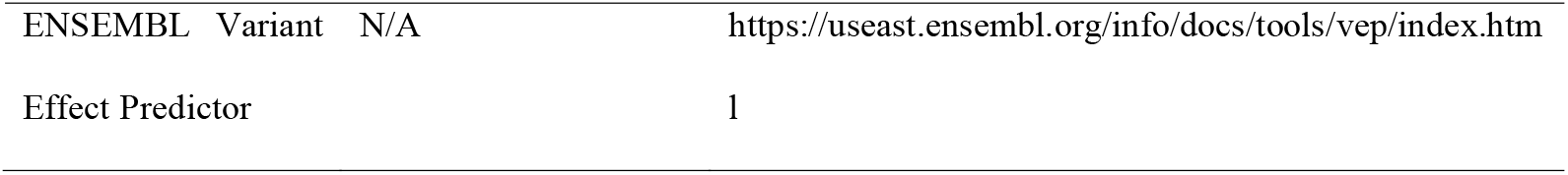

### LEAD CONTACT AND MATERIALS AVAILABILITY

#### Lead Contact

Further information and requests for resources and reagents should be directed to and will be fulfilled by the Lead Contact, Jia Qu.

### MATERIALS AVAILABILITY

This study did not generate new unique reagents.

### EXPERIMENTAL MODEL AND SUBJECT DETAILS

#### Overview of the High Myopia Sequencing Consortium cohort

The Myopia Associated Genetics and Intervention Consortium (MAGIC) is a large-scale genomic consortium integrating myopia cohorts and sequencing data from many investigators. Over the past several years, MAGIC has been able to collected samples at the Eye Hospital of Wenzhou Medical University (Zhejiang Eye Hospital) through the Institute of Biomedical Big Data (Xu et al., 2021). We recruited approximately ten thousand Chinese schoolchildren with high myopia aged from 6 to 18 from MAGIC. The analysis presented here is based on 21,227 unique human samples collected from epidemiological studies of myopia.

#### Informed consent and study approval

Research performed on samples and data of human origin was conducted according to protocols approved by the institutional review boards of the Eye Hospital of Wenzhou Medical University, and informed consent was obtained from all subjects.

### METHOD DETAILS

#### Whole-exome sequencing

A total of 21,227 MAGIC samples were sequenced on Illumina NovaSeq 6000 sequencers at Berry Genomics using the Twist Human Core Exome Kit. Sequencing reads were aligned to human genome build 37 (GRCh37) using the Burrows–Wheeler Aligner (BWA 0.7.12) (Li and Durbin, 2009), and Sambamba 0.6.6 (https://lomereiter.github.io/sambamba/) was used for sorting by chromosome coordinates and marking duplicates. Single-nucleotide variants (SNVs) and insertions/deletions (indels) were then called using a series of Genome Analysis Toolkit (GATK 4.0.11.0) commands: BaseRecalibrator, ApplyBQSR, HaplotypeCaller, CombineGVCFs and GenotypeGVCFs (Van der Auwera et al., 2013). Variant calling accuracy was estimated using the GATK Variant Quality Score Recalibration (VQSR) approach.

### QUANTIFICATION AND STATISTICAL ANALYSIS

#### Quality Control

##### Sample quality control and variant quality control

Variants were prefiltered so that only those passing the GATK VQSR (variant quality score recalibration) metric and those located outside of low-complexity regions were remained. Genotypes with a genotype depth (DP) < 10 and genotype quality (GQ) < 20 and heterozygous genotype calls with an allele balance >0.8 or <0.2 were set as missing. We excluded variants with a call rate < 0.9, a case–control call rate difference > 0.005, or a Hardy-Weinberg equilibrium (HWE) test *P* value < 10^−6^ on the basis of the combined case–control cohort. Samples were excluded if they showed a low average call rate (<0.9), low mean sequencing depth (<10), or low mean genotype quality (<65). Outliers (>4 SD from the mean) of the transition/transversion ratio, heterozygous/homozygous ratio, or insertion/deletion ratio within each cohort were further discarded. The number of samples and the variant dropout in each step were detailed in Tables S1 and S2.

##### Sex discrepancy

Samples with an X chromosome inbreeding coefficient > 0.8 were classified as males, while samples with an X chromosome inbreeding coefficient < 0.4 were classified as females. Samples between <0.8 and >0.4 which classified as ambiguous sex status, were excluded from the dataset.

##### PCA

PCA was performed using PLINK 1.07. We used a subset of high-confidence single-nucleotide polymorphisms (MAF>1%) in the exome capture region to calculate the PCs. A series of principal component analyses (PCAs) were performed to identify ancestral backgrounds and control for population stratification. Only retained individuals of East Asian (EAS) ancestry were retained, which were classified by a random forest algorithm with 1000 Genomes data (Figure S2, Table S1). Within the EAS population, we removed the controls that were not well matched with cases on the basis of the top three PCs by PCAmatchR (Brown et al., 2021)(Figure S3, Table S1).

##### Relatedness check

We included only unrelated individuals (identity by descent proportion < 0.2) using PLINK 1.07 (Figure S4, Table S1).

#### Variant harmonization

To alleviate confounding caused by differential call rates, we used a site-based pruning strategy similar to a previously described exon-pruning strategy was employed (Povysil et al., 2019). Individuals showing targeting of exonic sequence sites were excluded from analysis if the absolute difference in the percentage of cases compared with controls with an adequate call rate for the site differed by more than 0.007 (Figure S5). According to site-based pruning, 2.42% of the target exonic sequence bases were excluded from the respective analyses to alleviate issues related to differential call rates (Table S2). Furthermore, we removed 2.36% of the target exonic sequence bases that reached the genome-wide significance threshold (*P*<1×10^−6^) in the call rate association test (two-sided P value from Fisher’s exact test) (Figure S6). The advantage of this pruning was that the opportunity to detect variants was harmonized between the sequenced case and control samples, thereby reducing any bias in the number of detected variants that were not involved in disease risk (Figure S7).

#### Variant annotation

The annotation of variants was performed with Ensembl’s Variant Effect Predictor (VEP v.99) for human genome assembly GRCh37. We used the VEP (McLaren et al., 2016) CADD, LOFTEE (https://github.com/konradjk/loftee) and SpliceAI plugins to generate additional bioinformatic predictions of variant deleteriousness. Protein-coding variants were annotated into the following four classes: (1) synonymous; (2) benign missense; (3) damaging missense; and (4) protein-truncating variants (PTVs). In detail, using VEP annotations (v.99), missense variants were classified as “inframe_deletion”, “inframe_ insertion”, “missense_variant” or “stop_lost” variants. Among the missense variants, one type of benign missense variant was predicted as “tolerated” and “benign” by PolyPhen-2 and SIFT, respectively, and another type of benign mutation showed a combined annotation dependent depletion (CADD) score < 15. Furthermore, damaging missense variants were predicted as “probably damaging” and “deleterious” by PolyPhen-2 and SIFT and CADD > 15. Finally, PTVs were classified as “frameshift_variant”, “splice_acceptor_variant”, “splice_donor_ variant”, “stop_gained”, or “start_lost” variants, and variants were labeled as “high-confidence” (HC) according to LOFTEE and SpliceAI scores larger than 0.5 in the predictions of SpliceAI (Table S7). Allele frequencies were estimated among our case–control samples from three external exome sequence databases (1000 Genomes, ESP and gnomAD). We classified variant allele frequencies using the following criteria: (1) common (MAF > 5%); (2) low-frequency (0.5%<MAF<5%) and (3) rare (MAF < 0.5%) (Table S3).

#### Association Test

##### Exome-wide single-variant association analysis

We conducted two types of single-variant association analyses: the (two-sided) MLMA (Yang et al., 2011), MLMA-LOCO (Yang et al., 2011), BOLT-LMM (Loh et al., 2015) and EMMAX tesst (Kang et al., 2010) were used for all (including related) samples, while the (two-sided) Firth logistic regression test (Flannick et al., 2019) was used for unrelated samples. The Firth analysis included covariates of principal components of genetic ancestry (those among the first 10 PCs). We used the MLMA-LOCO results for association *P* values and the Firth results for effect size estimation. The test statistics obtained via linear regression were inflated because of the population differentiation caused by genetic drift. Post hoc correction approaches, such as “Genomic Control”, were used to correct the inflation (Devlin and Roeder, 1999).

For the exome-wide association study (ExWAS), we first tested each variant, regardless of allele frequency, for HM associations; we applied a significance level of *P* < 4.3 × 10^−7^ for all variants (Sveinbjornsson et al., 2016). For an MAF > 0.05, approximately 80% power was used to detect variants with a multiplicative genotype relative risk of 1.23 (Figure S8). To estimate cross-population genetic-effect correlations between Chinese in MAGIC and European in UK Biobank, we used Popcorn (V1.0) software (Brown et al., 2016) and *r*_b_ (Qi et al., 2018), the summary data-based approaches for estimating cross-population genetic correlation. We performed meta-analysis of summary-level ExWAS results using an inverse-variance-weighted fixed-effects model with METAL software (Willer et al., 2010).

##### Exome-wide burden analysis

To determine whether the enrichment of a specific class of variation was present in HM cases versus controls, we applied multiple Firth logistic regression models. Model 1 predicted HM case–control status solely from the variant count; Model 2 incorporated multiple covariates (sample sex and sample population structure from the first ten PCs); Model 3 incorporated all covariates used in the second model along with the sample total exome count, which is the exome-wide count of variants in the specific frequency class tested. The three Firth logistic regression models were used to predict case–control status from the exome-wide counts of synonymous, benign missense, damaging missense and PTVs. We performed an enrichment analysis of genes associated with myopia in HM cases using Firth logistic regression models. Model 3, considered to be the most conservative one representing the dataset, was used as the preferred model for our analysis. The myopia-related genes and genes involved in retinal blood vessel morphogenesis and neurotransmitter transport were listed in Table S8.

##### Gene-Based Collapsing Analysis

To determine whether a single gene was enriched in or depleted of rare protein-coding variants in HM cases, we performed four gene-level association tests including Fisher’s exact test, burden, SKAT and SKAT-O, with previously defined covariates (sample sex, PC1-PC10). The threshold of exome-wide correction for multiple testing was set as *P* < 3.66 × 10^−6^ (0.05/13,649 genes), which was calculated as the 5% type I error rate divided by the number of genes. We performed the following two tests for HM cases and controls: one test for rare PTVs and another test for rare damaging missense variants. To account for population stratification in our rare variant association study, we also applied SAIGE-GENE (Zhou et al., 2020) to analyze protein-coding genes for HM case-control associations in our cohorts, which showed substantial sample relatedness.

#### Expression Analysis

##### Tissue-specific expression

The gene-level reads per kilobase million mapped reads (RPKM) values of the Genotype-Tissue Expression (GTEx) RNA-seq data (https://www.gtexportal.org/home/) were used across 55 tissue types, including peripheral retina samples(Lonsdale et al., 2013). Genes were defined as eye-expressed genes if they were present with an RPKM of 0.5 in 80% of the samples from at least one tissue type. The expression values were log-transformed (log2[RPKM+1]). To determine tissue-specific gene expression signatures, a linear regression model was applied for each gene in each tissue against all other tissues. Models were adjusted for age, RIN, sex, individual as a repeated-measure, and surrogate variables to account for potential batch effects and other unwanted technical and biological variation. Significance values were adjusted for multiple testing using the Benjamini and Hochberg (BH) method to estimate FDRs. After the BH correction, genes with q value < 0.05 and T value > 3 were defined as “tissue-specific” in a given tissue, especially if the tissues were closely related.

##### Cell type enrichment

Single-cell expression profiles from the adult peripheral and foveal retina were downloaded from Mendeley Data (https://doi.org/10.17632/sm67hr5bpm.1) (Cowan et al., 2020) and used to identify the cell-type specificity of candidate genes. Single-cell expression values were log-transformed and centered using the mean expression values. The average centered expression values of candidate genes were calculated for each cell. Cells were grouped into cell clusters (cone, rod, ganglion cell, retinal pigment epithelial cell, etc.), and the relative expression level of a given cell cluster was calculated by a scale function in R.

##### Weighted gene coexpression network analysis (WGCNA)

Weighted gene coexpression network analysis (WGCNA) (Langfelder and Horvath, 2008) was used to build a signed coexpression network for all peripheral retina samples. To construct a weighted gene network, the power (*b*) was selected using the scale-free topology criterion previously outlined by Zhang and Horvath (Zhang and Horvath, 2005). The setting of *b* = 8 was applied. A signed network construction method was used to identify coexpression modules using the blockwiseModules function. We chose a relatively large minimum module size of 30, a threshold for merging modules of 0.2 and a medium sensitivity (deepSplit=2) for cluster splitting.

The strength of interaction (correlation) between each module and the intramodular connectivity of genes in the corresponding modules of interest was measured using module eigengene-based connectivity (kME). The genes with the highest kME (|kME|>0.7), which represented the central status in the module gene network, were selected as the hub genes in the corresponding module and used for enrichment analysis. WGCNA yielded 12 retinal coexpression modules, one of which showed significant overrepresentation of 32 HM genes, 110 RP genes and 10 RD genes, but not RE genes or RA genes, after correction for multiple testing.

#### ChIP-Seq and ATAC-Seq Analysis Pipeline

ChIP-Seq and ATAC-Seq raw reads downloaded from GEO (GSE95632 and GSE137311) were aligned to the human reference genome sequence version GRCh37 using Bowtie2 (version 2.3.4) (Langmead and Salzberg, 2012). Quality control of the aligned BAM files was performed with SAMtools (Li et al., 2009), enabling only uniquely mapped reads to be retained, and PCR duplicates were removed with Picard (“Picard Toolkit” 2019; Broad Institute, GitHub Repository; http://broadinstitute.github.io/picard/; Broad Institute) for subsequent analyses. Significant H3K27ac and ATAC-seq peaks were called using MACS2 with all default parameters except for -p 1e-9 and -f BAMPE (Zhang et al., 2008). We then used the deepTools (Ramírez et al., 2016) bamCompare function to calculate the ChIP-Seq signal, subtract the corresponding background input signal, normalize the number of reads per bin under the RPKM method and generate BigWig format files. The signal intensities of each ChIP library were scored against the corresponding input library. The input-subtracted peak signal within a region was measured as the RPKM value using bigWigAverageOverBed.

#### Motif analysis

To detect sequence motifs associated with a significant association variant (rs533280354), we performed motif analysis of the 200 bp regions upstream and downstream of variant sites using HOMER (Heinz et al., 2010) with the default parameters except for -size given and -mask.

#### Functional enrichment analysis

Metascape (Zhou et al., 2019) was used for gene ontology and pathway analysis. Each term presented a P value (accumulative hypergeometric distribution) < 0.01 and an enrichment factor > 1.5 (the enrichment factor is the ratio between the observed counts and the counts expected by chance). Enrichment analysis were performed using Enrichr (Kuleshov et al., 2016) for the module genes.

#### Random gene selection

For top 50 HM risk genes that were significantly enriched in a retinal co-expression module, we also randomly sampled the same number of risk genes in the SAIGE-GENE results. We compared the ORs and *P*-values of HM risk genes enriched for module genes with the simulated ORs and *P*-values of randomly selected genes. This process was repeated for 1,000 times, and a empirically permuted *P*-value was calculated to test whether the HM risk genes showed significant enrichment of expression in the retina over that expected by random chance overall.

#### Luciferase reporter assay

The wild-type and mutant FKBP5 promoters were fully synthesized by BGI (The Beijing Genomics Institute) and inserted into the pGL3-basic backbone using the HindIII restriction site. siRNA was synthesized by Ribobio. siRNA duplexes targeting KLF15 mRNA or siNC were transfected into HEK293 cells using (https://www.thermofisher.com/us/en/home/brands/product-brand/lipofectamine/lipofectamine-rnaimx.html) Lipofectamine RNAiMAX transfection reagent (Invitrogen) as described in the manufacturer’s instructions. After a 48-h incubation, pGL3-WT or pGL3-MUT was cotransfected into cells with pRL-TK using Lipofectamine 2000 (Invitrogen). After an additional 24-h incubation, dual-luciferase activity was measured using a Dual-Glo Luciferase Assay kit (Promega). A t test was used to compare the fluorescence intensity among experimental groups.

## Notes

### Competing Interest Statement

The authors have declared no competing interest.

### Clinical Trial

ChiCTR2000041373

