## Supplemental Figures for "Sequencing of 19,219 exomes identifies a low-frequency variant in *FKBP5* promoter predisposing to high myopia in a Han Chinese population"

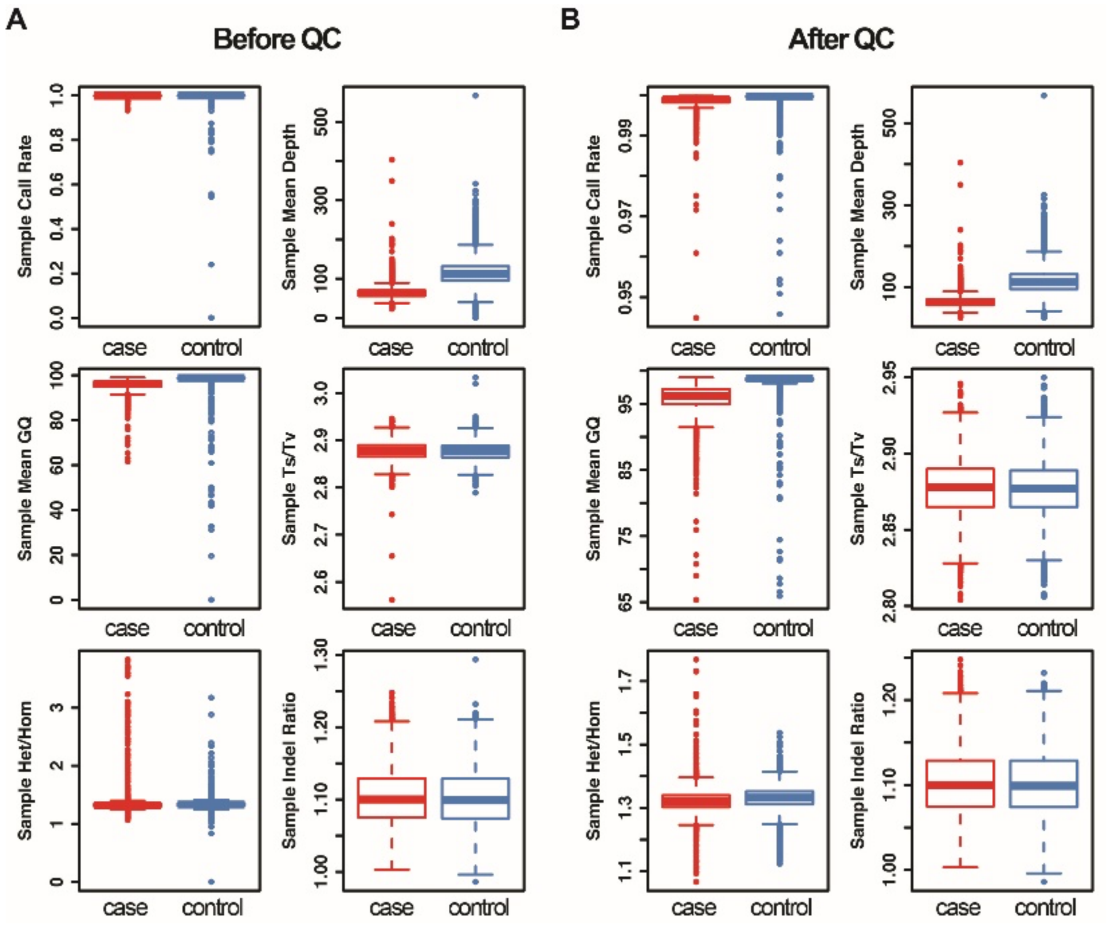


**Figure S1. Initial sample quality control analysis.** Distribution of sample call rate, sample mean depth, sample mean genotype quality, sample transition to transversion ratio, sample heterozygous to homozygous ratio and sample insertion to deletion ratio before (A) and after quality control (B). The box and whisker plots display the mean, minimum, and maximum.

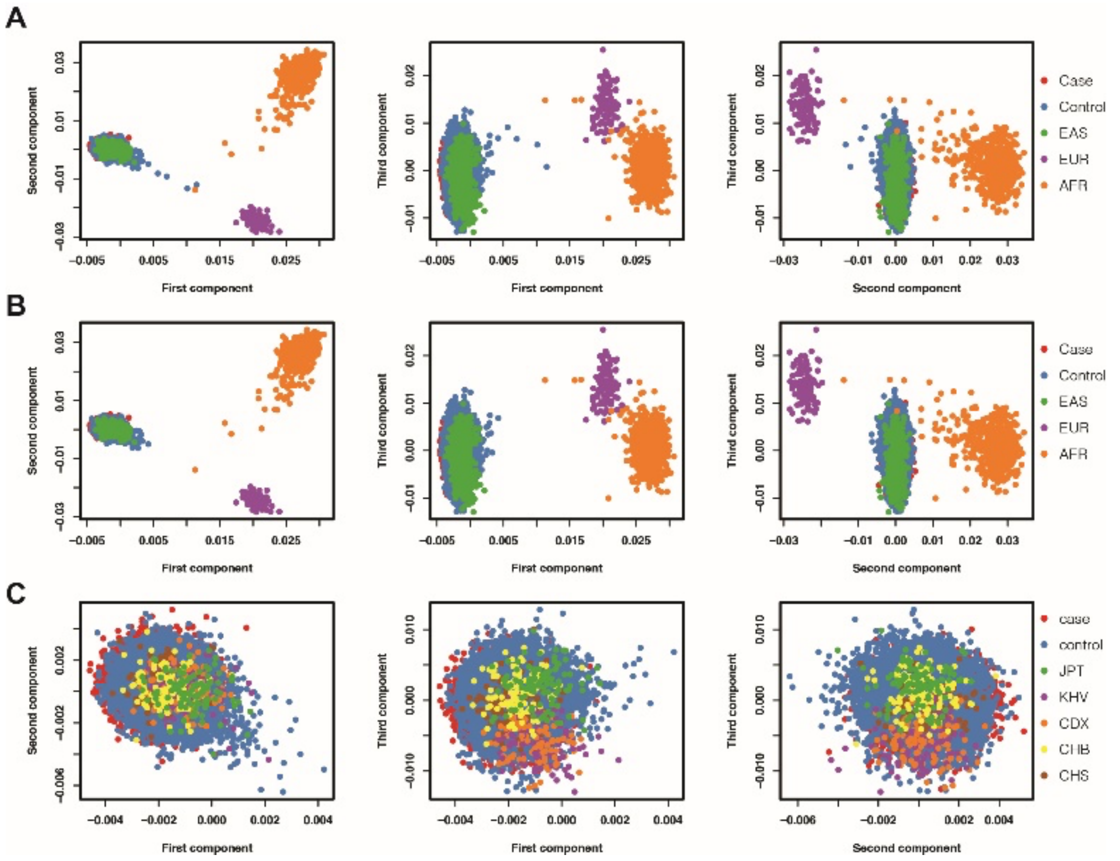


**Figure S2. Principal component analysis of MAGIC dataset with 1000 Genomes.** (A) PCA was run on the study samples along with 1000 Genomes (1KG) phase 3 super populations to infer genetic ancestry. HM cases and control samples were mostly of East Asian descent. (B) Genetic ancestry of the HM cases and controls were predicted by the Random Forest classifier based on top 6 PCs, using 1KG samples as the training data. Individuals were assigned to a particular 1KG-ancestry with a predicted probability >0.9, as depicted in the figure. We removed samples with unknown or unclassified ancestry and focused on East Asian samples-the largest population under study with abundant cases and controls-moving forward. (C) PC1, PC2 and PC3 of HM cases and controls with the East Asian subpopulations of 1000 Genomes.

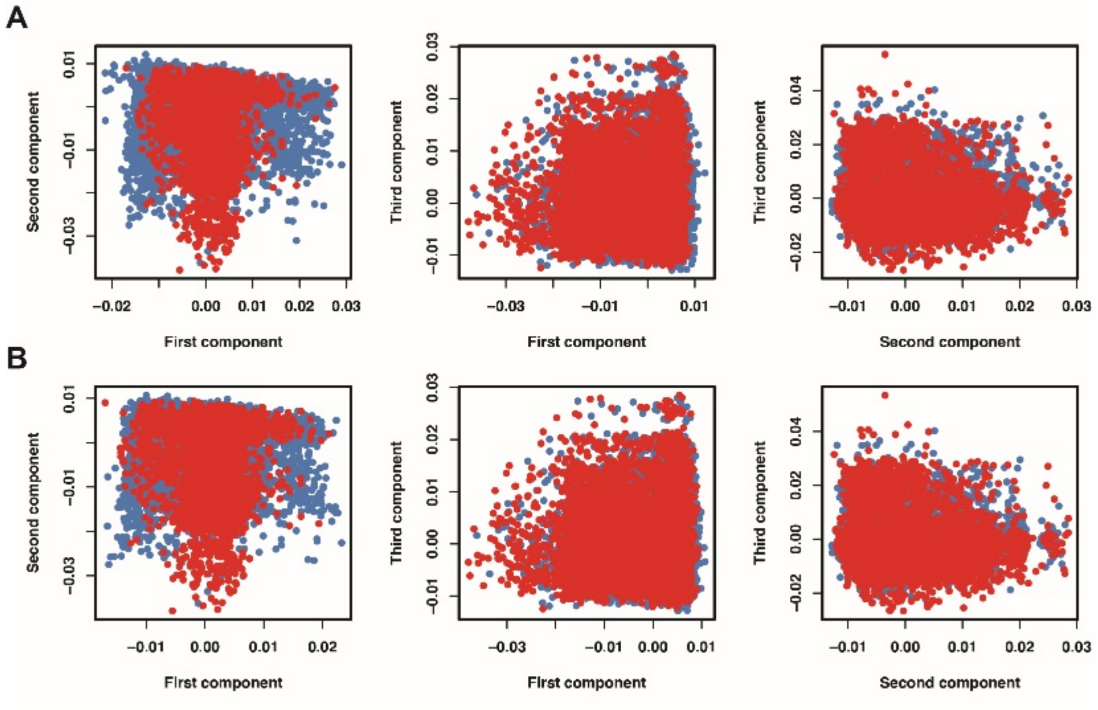


**Figure S3. PCA on MAGIC samples.** (A) Initial PCA on individuals classified as having an East Asian descent. Ancestry was not well-matched between cases and controls. (B) PCA after removing controls not pair-matched with cases based on top 3 PCs.

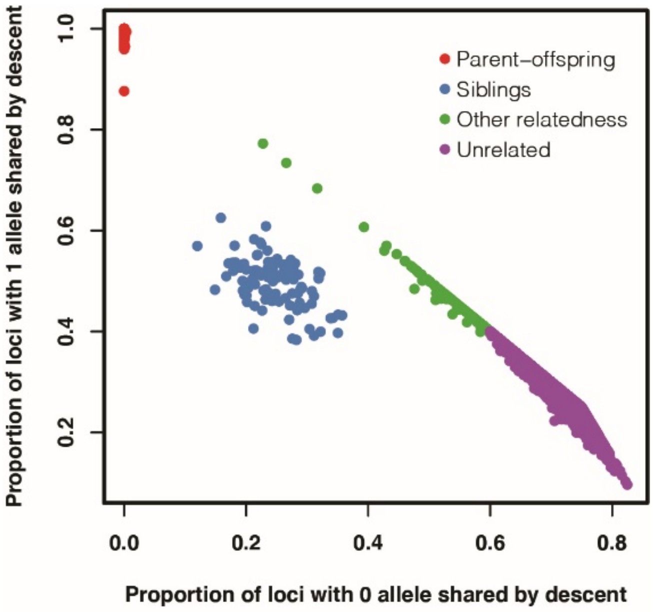


**Figure S4. Identify-by-descent (IBD) estimate for ancestry-matched (EAS) samples.** This plot shows only pairs of individuals with a relatedness > 0.125 for clarity. One individual in each of the related pairs with IBD > 0.2 was removed. Samples appearing in multiple related pairs were removed first, and cases were preserved over controls.

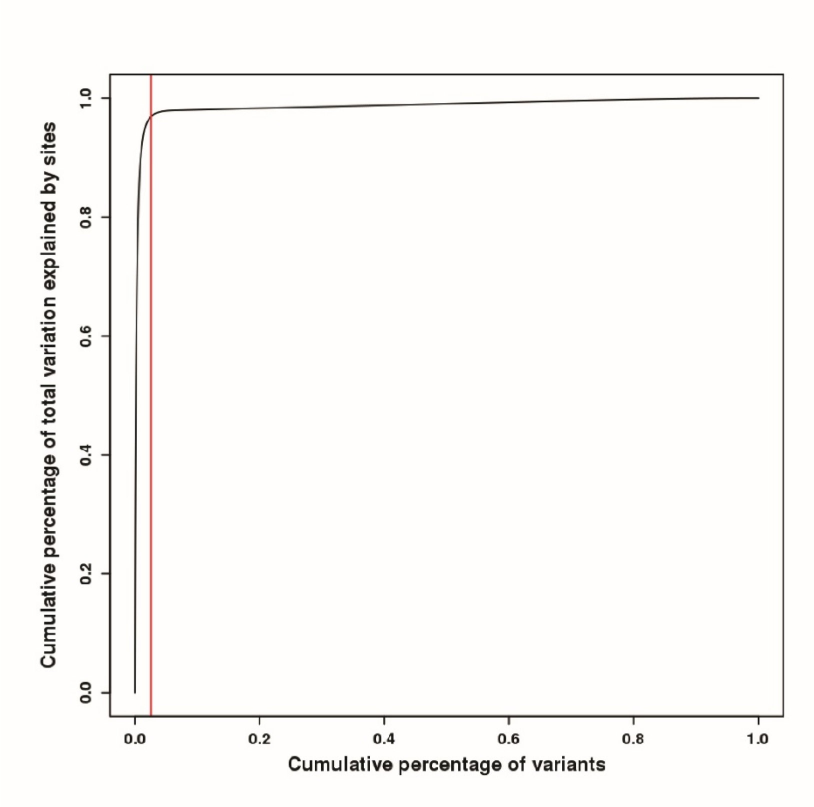


**Figure S5. Cumulative sum of variation plot for site pruning.** The y-axis reflects the cumulative sum of variation explained as sites with the largest variation in call rate are pruned out (x-axis). The red line represents the point at which we maximize the amount of study-wide variation explained while minimizing the percentage of the exome that is pruned out (2.6%). Identifying the exome site that occupies the 2.6% position translates back to a site that had difference 0.007 when comparing the HM case to the control cohort.

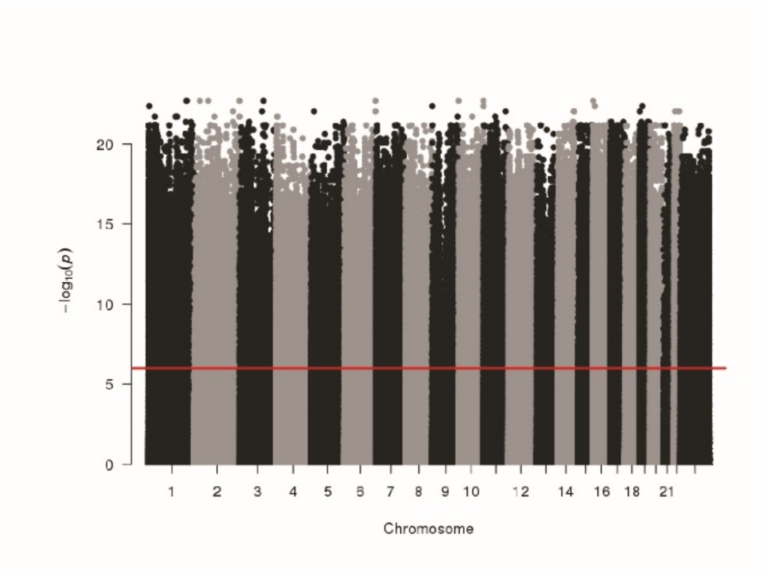


**Figure S6. Manhattan plot shows the association test of call rate between cases and controls by Fisher's Exact test.** Red line: exome-wide significance ($P<1\times10^{-6}$).

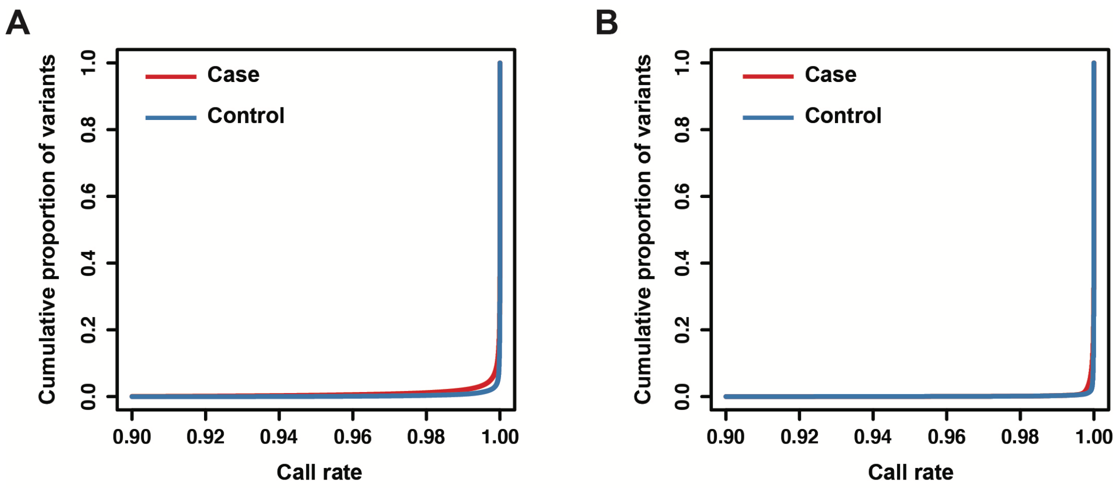


**Figure S7. Cumulative plot of call rate before (A) and after (B) variant harmonization.**

Red: case, Blue: control.

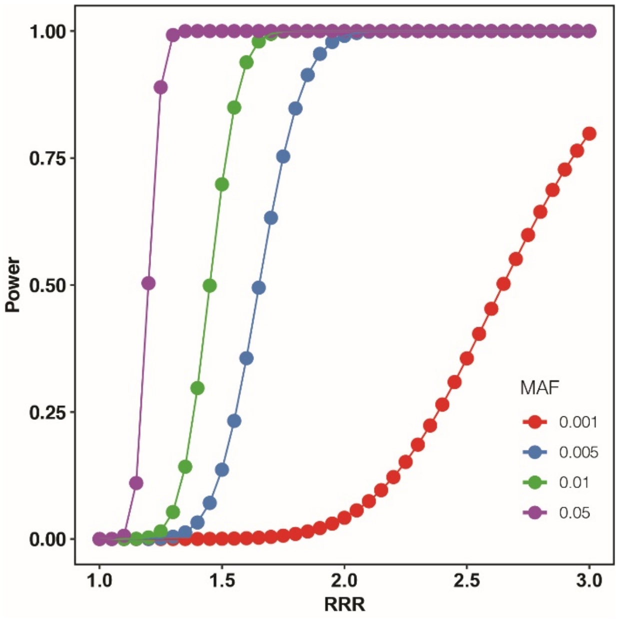


**Figure S8. Power analysis at a series of MAF threshold.**

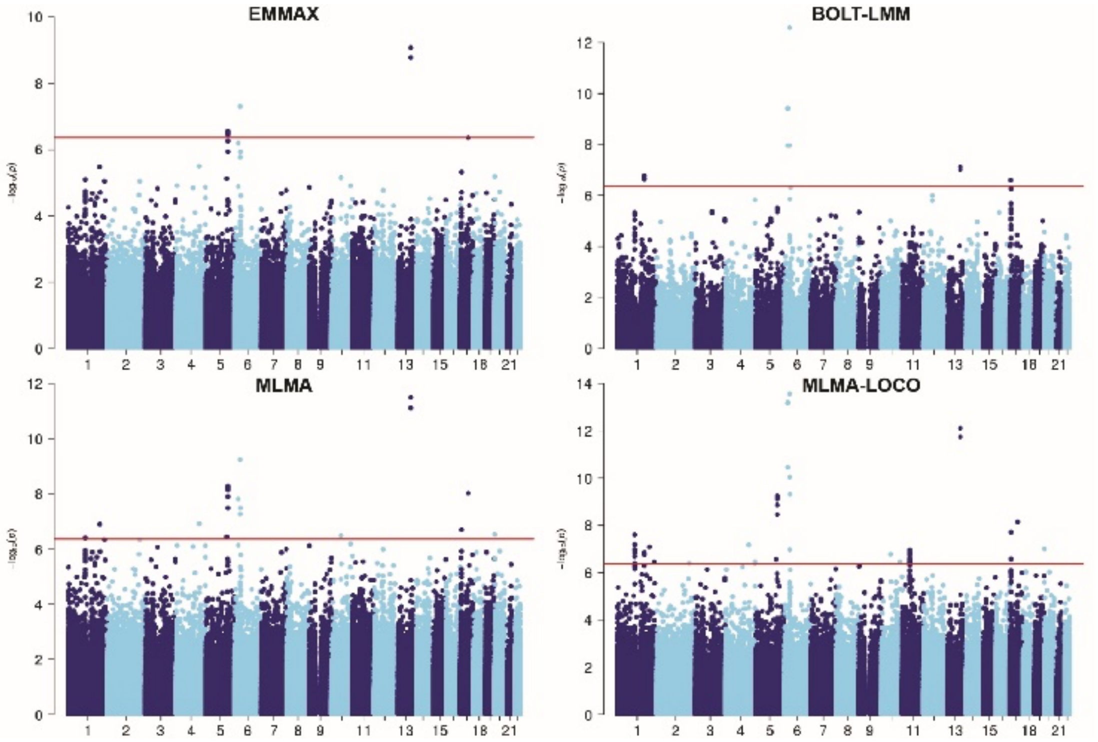


**Figure S9. Exome-wide association study for single variants by EMMAX, BOLT-LMM, MLMA and MLMA-LOCO.** Red line: exome-wide significance ($P < 4.3 \times 10^{-7}$).

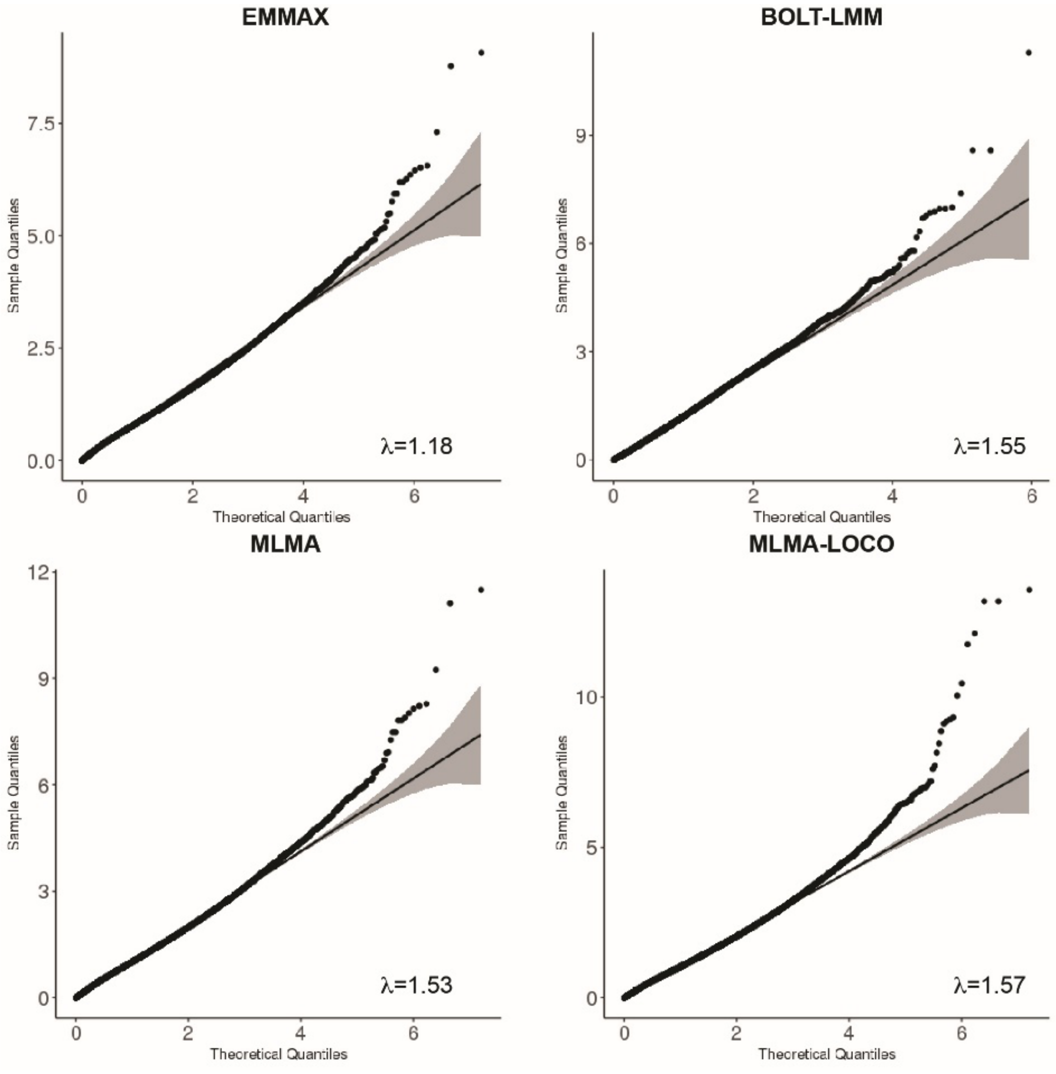


**Figure S10. QQ plots show moderate genomic inflation for exome-wide association studies**. Black line, expectation of P values under the null distribution. Grey region, 95% confidence interval of expectations under the null distribution.

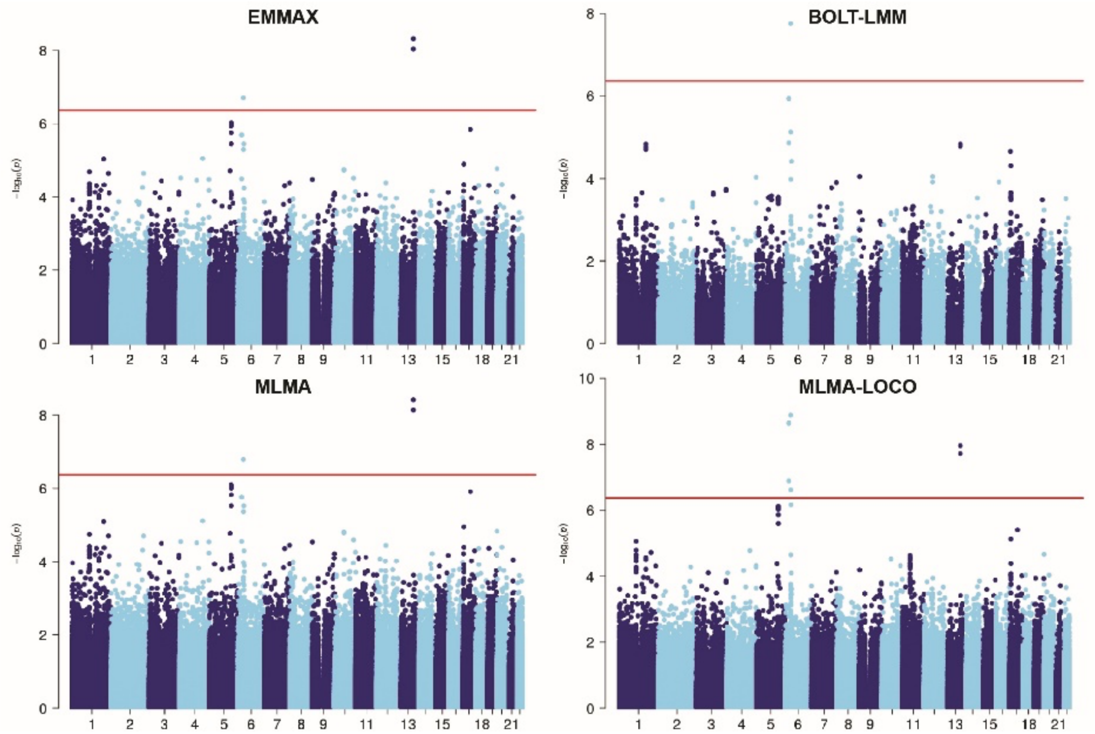


**Figure S11. Exome-wide association study for single variants by EMMAX, BOLT-LMM, MLMA and MLMA-LOCO.** Adjusting the test statistics by the genomic control (GC) inflation factor. Red line: exome-wide significance ($P < 4.3 \times 10^{-7}$).

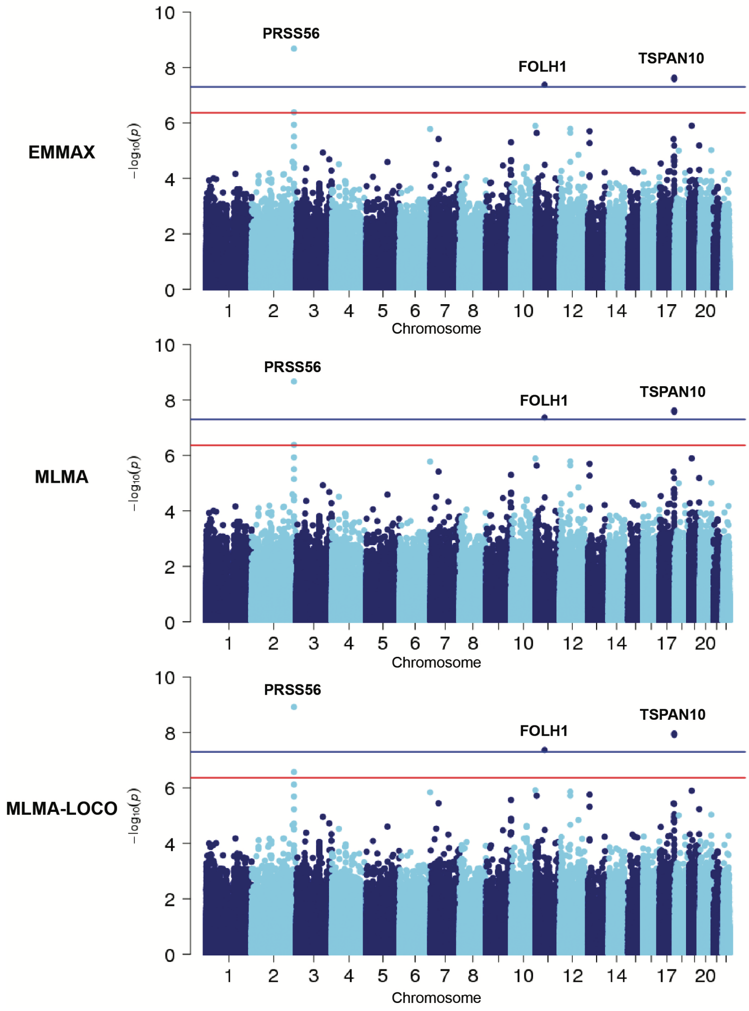


**Figure S12. Exome-wide association study for single variants by EMMAX, MLMA and MLMA-LOCO in UKB European cohort.** Adjusting the test statistics by the genomic control (GC) inflation factor. Red line: exome-wide significance ($P < 4.3 \times 10^{-7}$). Blue line, $P = 5 \times 10^{-8}$.

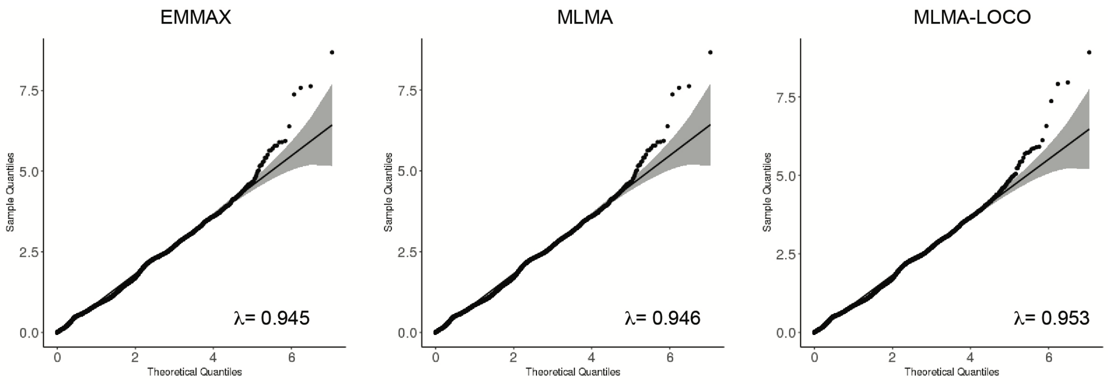


**Figure S13. The quantile–quantile plots of associations are shown for the *P*-value in UKB European cohort.** Black line, expectation of P values under the null distribution. Grey region, 95% confidence interval of expectations under the null distribution.

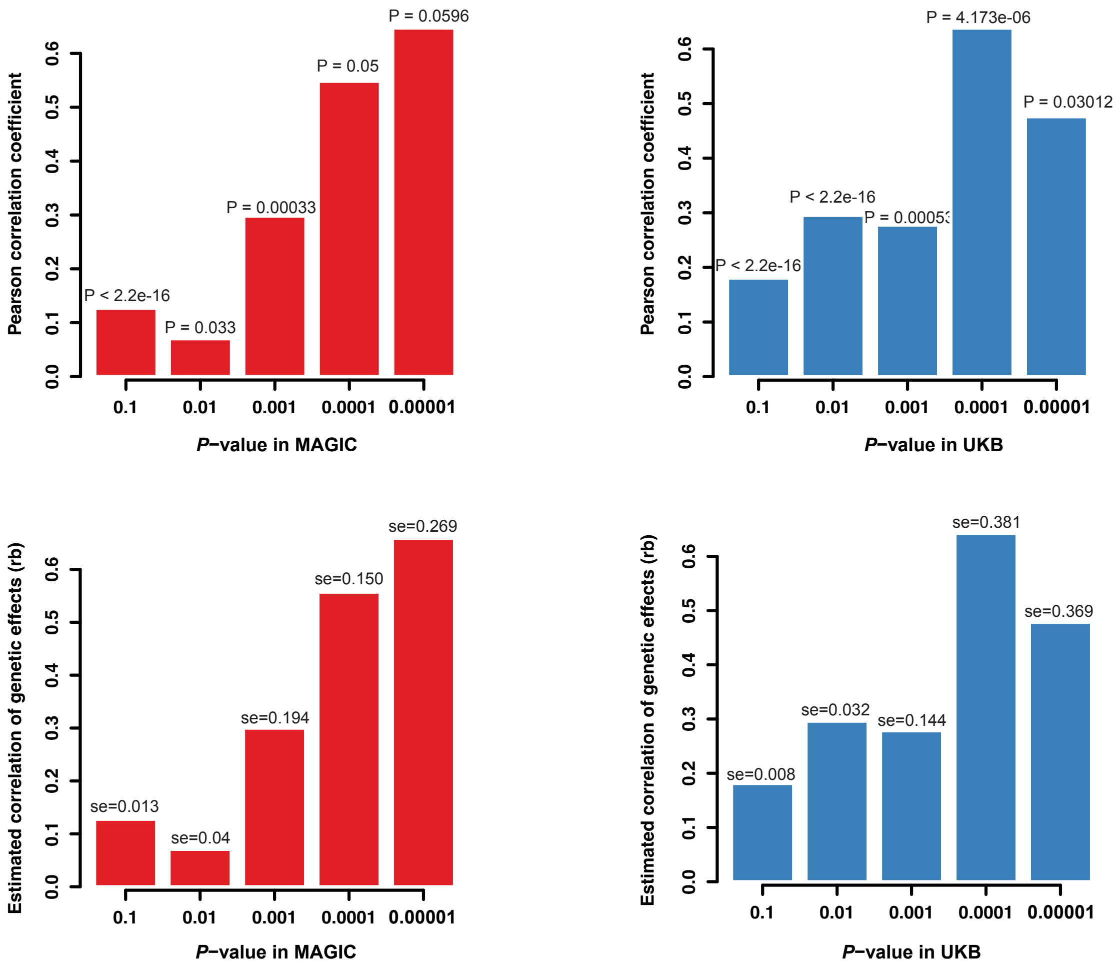


**Figure S14. Comparisons of the effect sizes (ORs) at different ExWAS P-value thresholds in the MAGIC (left) and UKB (right).** Bar plot displaying the Pearson correlation coefficient (up panel) and $r_b$ (bottom pane) of log(ORs) at P-value threshold. In the x-axis are reported the P-value thresholds used to filter for the variants to be included in the correlation computation.

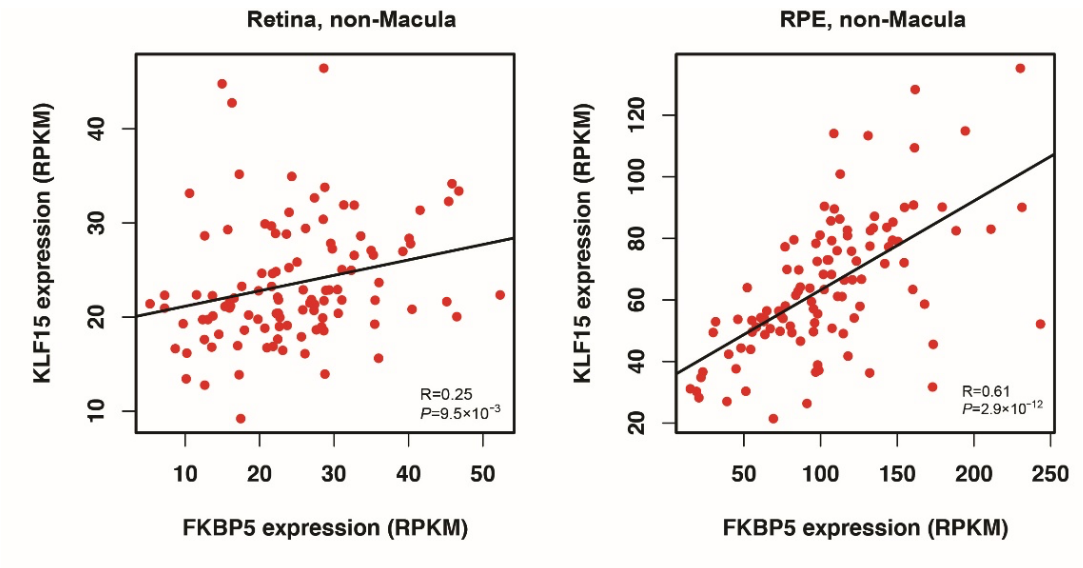


**Figure S15. Pearson correlations between FKBP5 and KLF15 expression in Retina and RPE of non-Macula, respectively.**

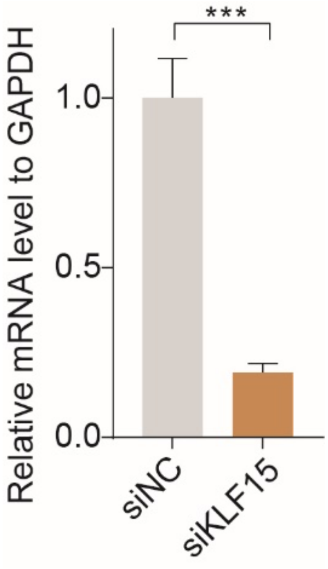


**Figure S16. siRNA inhibited KLF15 expression. KLF15 mRNA level of HEK293 cells following siKLF15 transfection, determined by RT-PCR.**

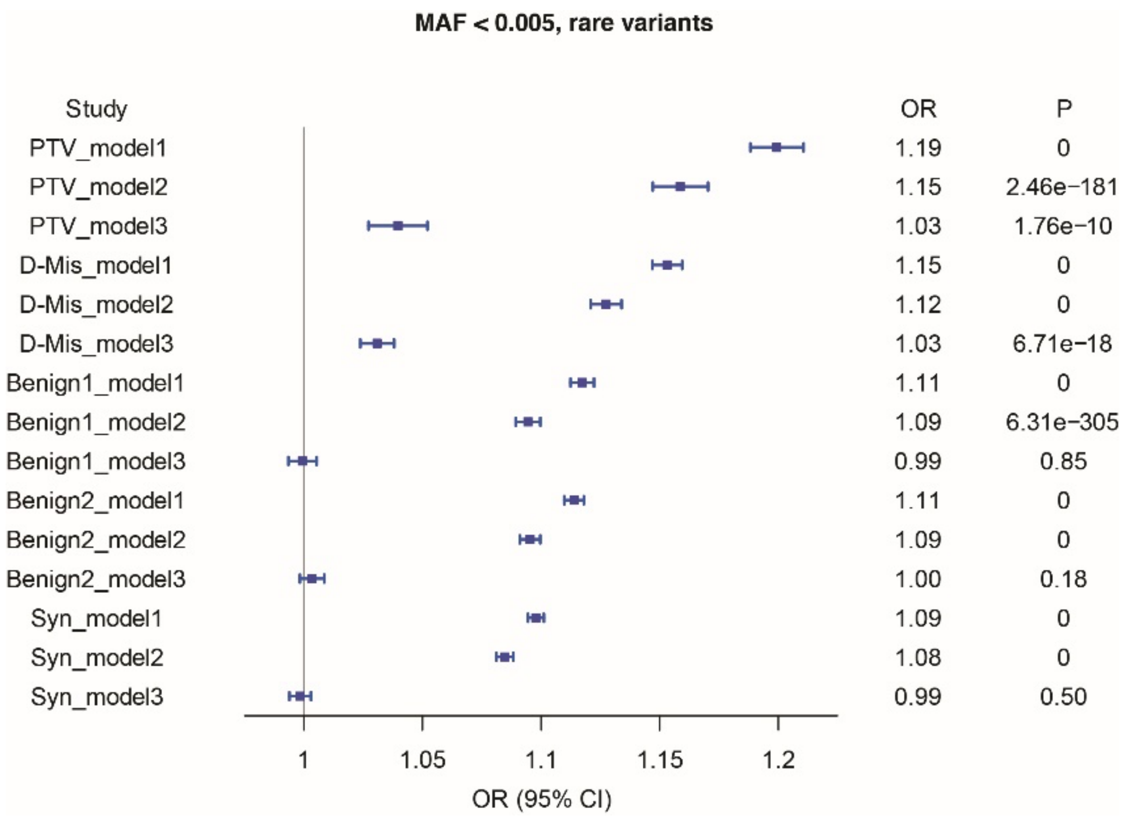


**Figure S17. Forest plot for burden analysis in variants that annotated within different types of protein-coding catalogues.**

Model 1: Sample variation. The graph displayed the mean and standard deviation. P-values from firth logistic regression test are also displayed.

Model 2: Sample variation, sample sex, PC1-PC10.

Model 3: Sample variation, sample sex, PC1-PC10, and total exome count (summation of synonymous, benign missense, damaging missense, and PTV).

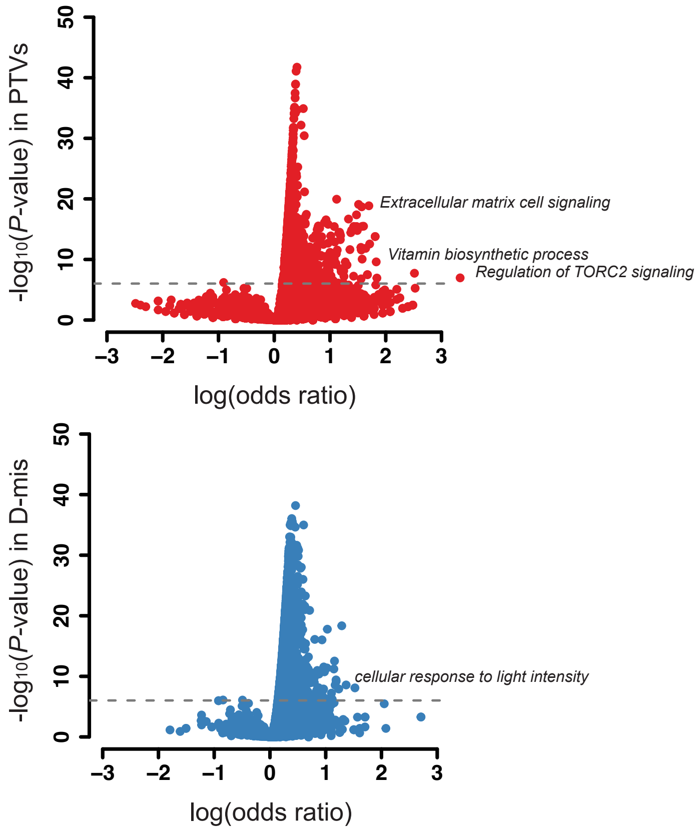


**Figure S18. Volcano plot of biological and empirical gene sets enrichment.** The pathway that enriched PTVs and D-mis after statistical analysis are reported in green and red. The horizontal line corresponds to a Bonferroni-corrected significance value of $< 0.05$.

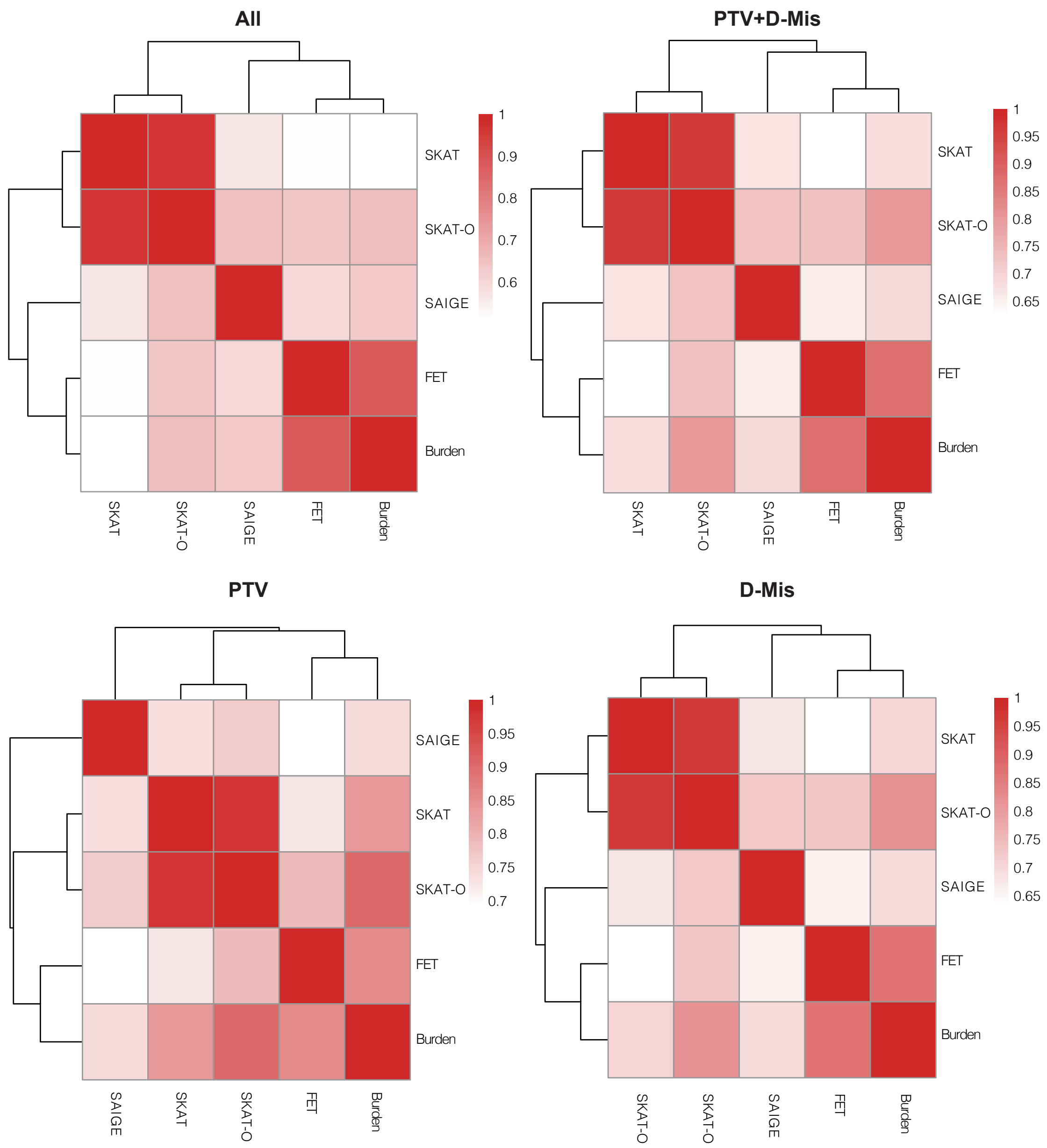


**Figure S19. Cross-correlation analysis of *P* values from five gene-level burden analysis methods.**

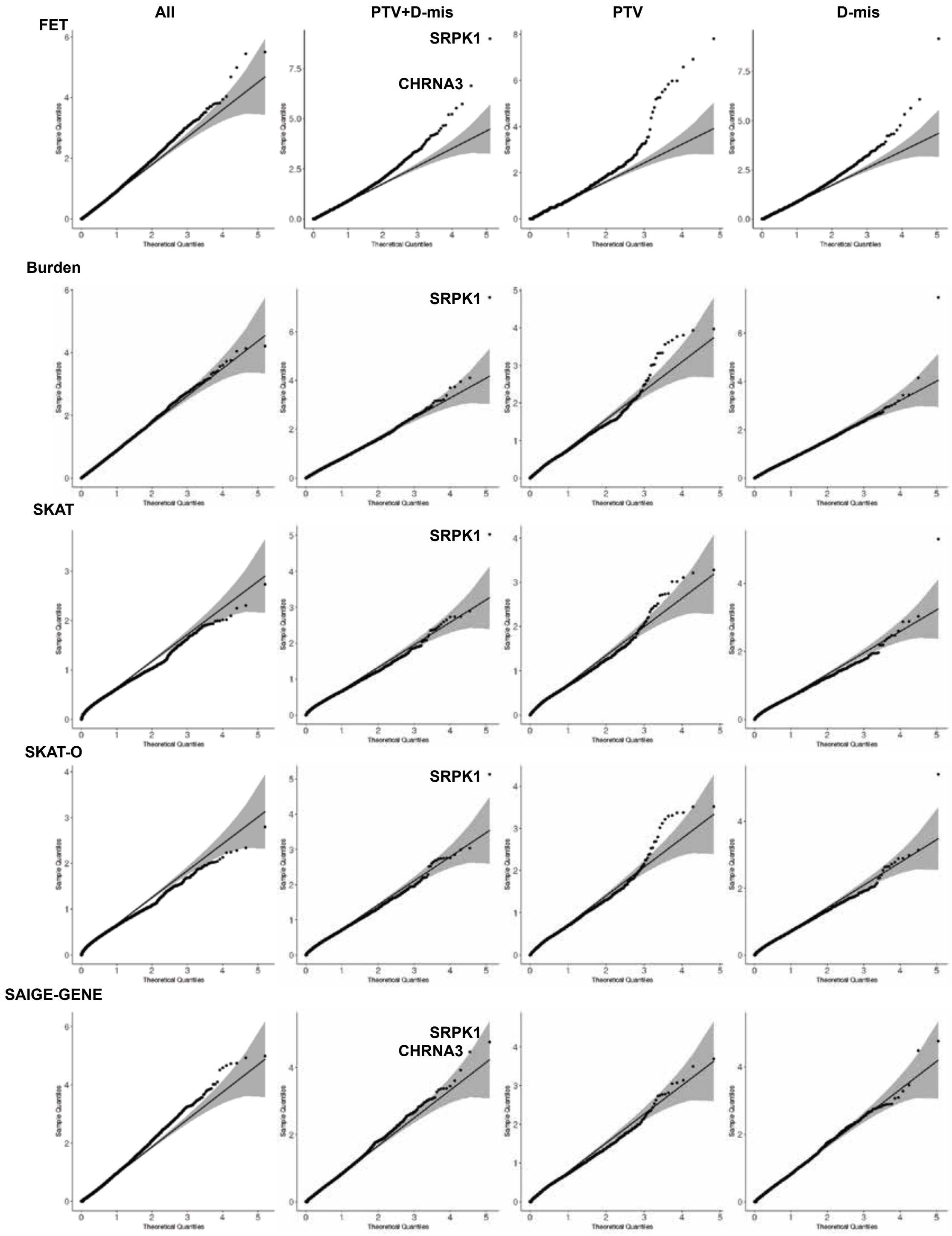


**Figure S20. QQ plots for gene-level burden analysis by FET, Burden, SKAT, SKAT-O and SAIGE-GENE under four models, respectively.**

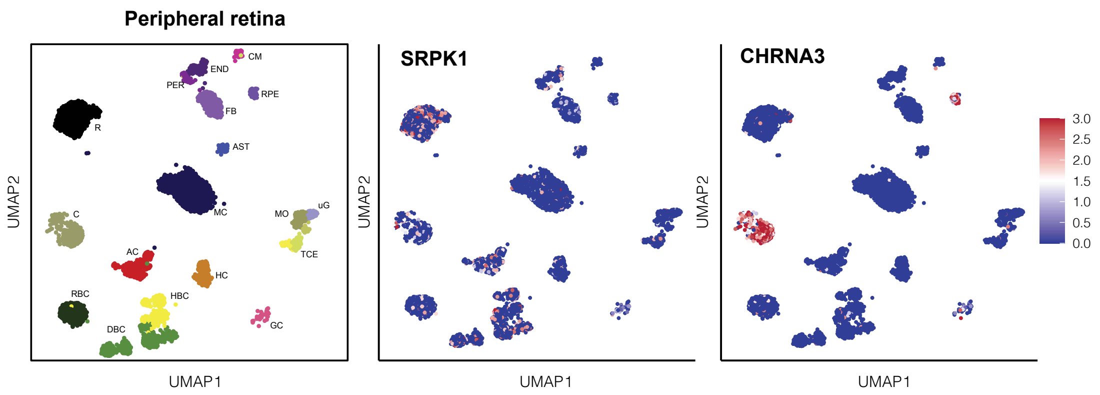


**Figure S21. UMAP of all tissues single-cell data with cell colored based on the expression of *SRPK1* and *CHRNA3* genes for particular cell types in peripheral retina.** Gene expression levels are indicated by shades of red.

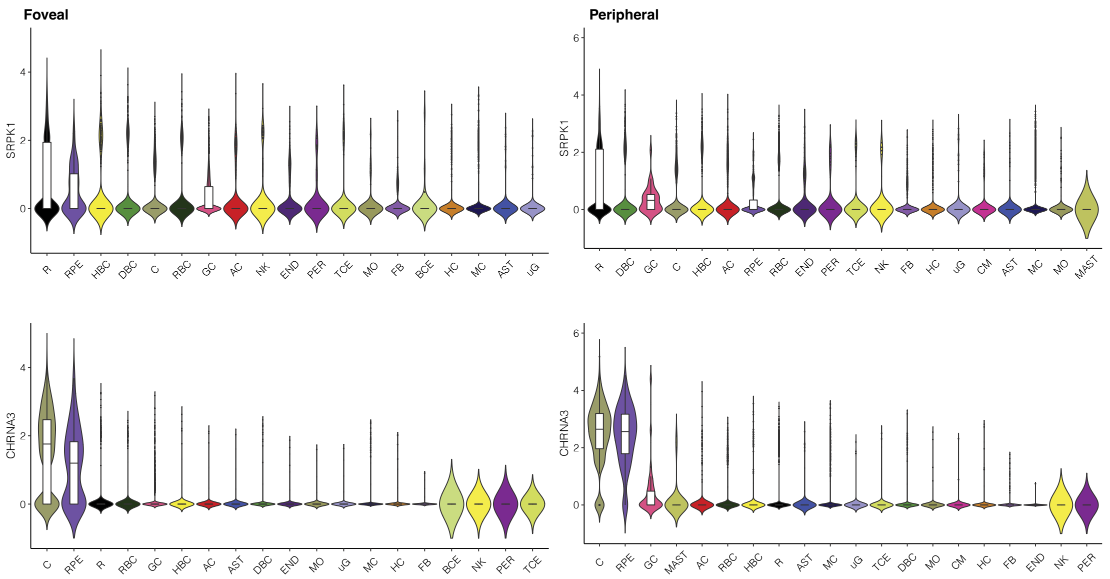


**Figure S22. Average expression level of *SRPK1* and *CHRNA3* in peripheral and foveal retina related cells.** C: cone; R: rod; RPE: retinal pigment epithelial cell; MC: Müller cell; GC: ganglion cell; BCE: B cell; AST: astrocyte; NK: natural killer cell; FB: fibroblast; AC: amacrine cell; PER: pericyte; uG: microglia; HBC: OFF bipolar; MO: monocyte; HC: horizontal cell; DBC: ON bipolar; RBC: rod bipolar cell; TCE: T cell; END: endothelial cell; CM: choroidal melanocyte; MAST: mast cell.

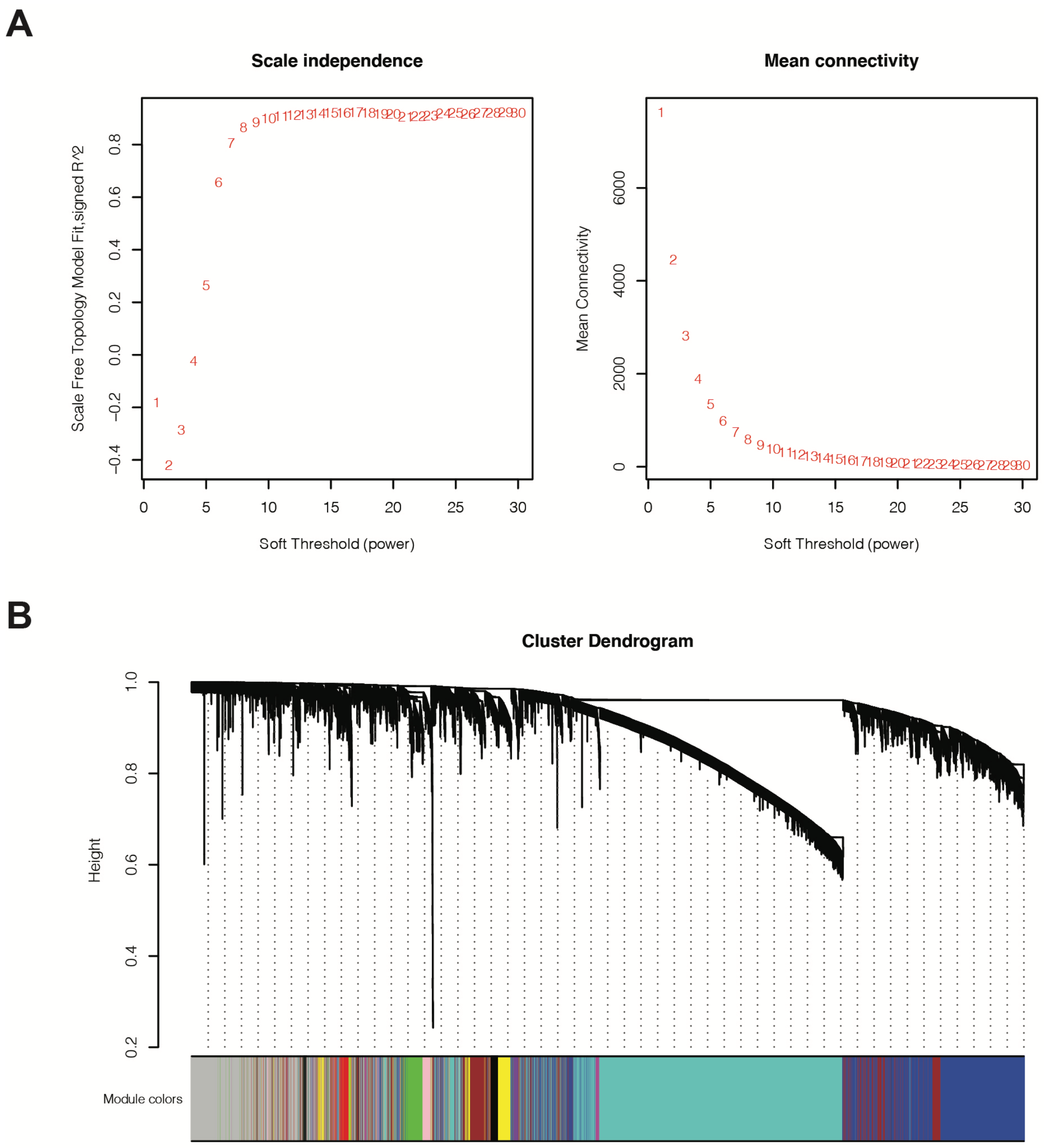


**Peripheral retina**

**Figure S23. Weighted gene co-expression network analysis all peripheral retina samples.** (A) power b threshold chosen by scale-free topology criterion; (B) Cluster dendrogram of co-expression modules.

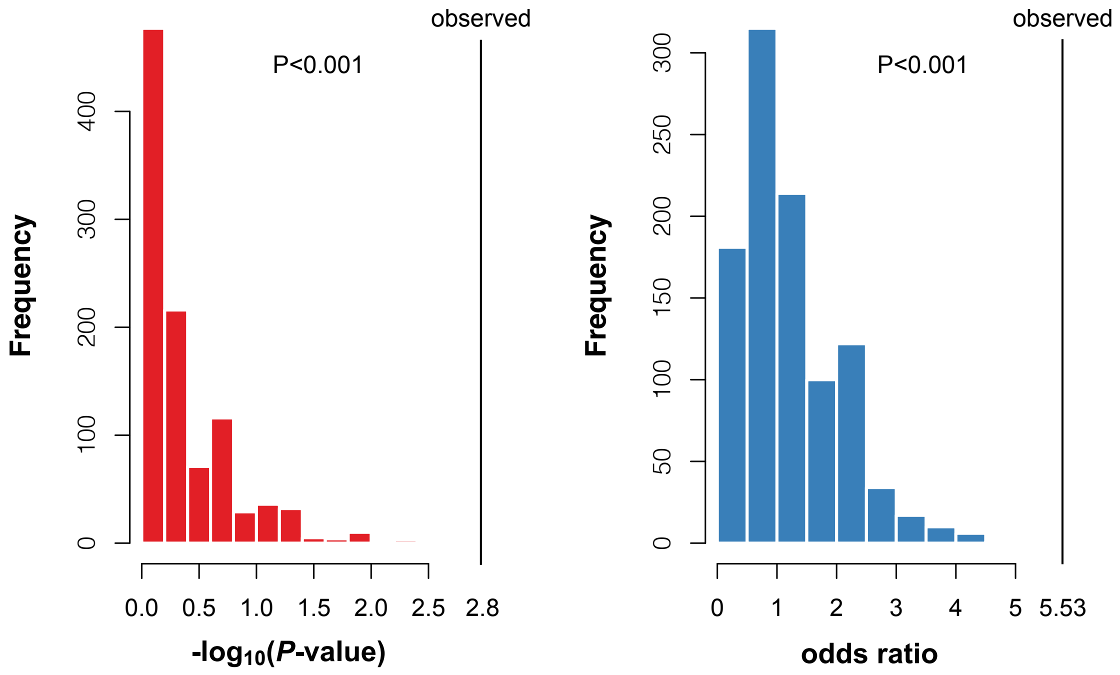


**Figure S24. Permutation test for HM genes in coexpression module**.The observed P-value and odds ratio of significant 50 HM risk genes enriched in a coexpression module "brown" versus random selection of 50 genes (the histogram, 1,000 replications of permutations were performed).
